# Resilience and Protective Factors for Mental Health among Indigenous Youth in Canada: A Scoping Review

**DOI:** 10.1101/2024.08.05.24311522

**Authors:** Hwayeon Danielle Shin, Leah Carrier, Jessy Dame, Michelle Padley, Anika Daclan, Helen Wong, Ronessa Dass, Rachel Anne Dorey, Emma Stirling-Cameron, Jodi Langley, Janet A. Curran

## Abstract

Indigenous youth’s inherent strength and resilience play a vital role in their well-being and mental health. Protective factors, closely linked to resilience, spanning individual, family, and community levels reinforce positive mental health outcomes. The purpose of the present scoping review was to summarize the available literature that describes resilience and/or protective factors promoting mental health and well-being among Indigenous youth in Canada. As a secondary objective, this review investigated community involvement reported in the identified sources. JBI scoping review methodology was followed, and the search of PubMed, EMBASE, CINHAL, PsycINFO, ERIC, and Scopus commenced in August 2021, and was updated in February 2023. A targeted Google search was also conducted to identify eligible grey literature. A total of 61 papers were included in data extraction. The types of sources identified were observational (n=22), participatory action research (n=11), mixed/multi-methods (n=10), qualitative (n=9), case study (n=4), quasi-experimental (n=1), experimental (=1), and other designs such as quality improvement and program evaluation (n=3). Additionally, only a handful of included studies reported use of an Indigenous-specific approach, such as Two-Eyed seeing. Protective and resilience factors were identified across various levels such as individual (n=52), interpersonal (n=37), and wider environmental beyond social systems (n=37) levels. Forty studies described community involvement, which included non-specified community members, like friends or citizens (n=21), youth (n=19), Indigenous community members such as leaders and workers (n=14), and Elders (n=11). These groups were engaged to varying degrees, functioning either as equal collaborators, consultants, or, in some instances, as decision-makers.

## Introduction

Indigenous peoples in Canada include First Nations, Inuit, and Métis, and there are specific communities within them, such as the Cree, Ojibwe, and Dene communities. Indigenous youth possess inherent strengths and resilience that foster health and well-being (Peters et al., 2013; Smylie et al., 2010). Indigenous communities describe resilience as a healing journey, enabling individuals to overcome the various traumas and cultural losses stemming from historical and colonization processes (Goulet et al., 2011; Isaak et al., 2015; Tousignant & Sioui, 2009). The significance of culture in promoting mental well-being and resilience within Indigenous and ethnic/racial minority communities have been well documented in the literature (Kirmayer et al., 2011; Ungar & Theron, 2020). Protective factors, which are tightly linked to resilience, exist across individual, family and community levels, and they also promote positive mental health outcomes (MacDonald et al., 2013). Thus, these protective and resilience factors, all of which are inherent strengths within Indigenous youth, represent powerful assets for mental health and well-being. Yet they often remain overshadowed by the emphasis on challenges, vulnerabilities, and risk factors in mental health research.

Over the years, several reviews have contributed to our understanding of resilience among Indigenous youth, highlighting its multifaceted nature and the vital role of cultural, communal, and personal elements. In 2017, Rowhani and Hatala emphasized the significance of cultural continuity, community bonds, and empowerment in promoting resilience (Rowhani & Hatala, 2017). In 2016, Toombs and colleagues highlighted that resilience combines individual, community and cultural elements that are crucial for fortifying strength and fostering endurance among Indigenous youth which led to positive mental health outcomes (Toombs et al., 2016). Most recently in 2022, Heid and colleagues also echoed that resilience is complex and dynamic process and that resilience strategies involved future orientation, cultural pride, learning from nature, and community interactions for Indigenous youth in both Canada and USA (Heid et al., 2022).

Expanding upon earlier research initiatives and incorporating insights from grey literature, the present scoping review seeks to identify and summarize available literature that describes resilience and/or protective factors for mental health and well-being among the Indigenous youth in Canada. Although mental health cannot be separated from overall well-being in Indigenous conceptualizations of health, there are specific mental health concerns that are particularly salient and need targeted attention, such as suicide and substance misuse (Canada, 2019; Sikorski et al., 2019). We aim to bring strengths to the forefront, shedding light on the inherent capacities Indigenous youth possess to navigate adversity and foster their mental health and wellness. Furthermore, this review aims to include a comprehensive range of evidence types, recognizing that Indigenous knowledge and ways of knowing are frequently documented in grey literature and personal narratives. This is particularly pertinent since Indigenous viewpoints on mental health can substantially diverge from Western perspectives.

As such, our review will incorporate diverse information sources typically omitted from traditional systematic reviews including those mentioned earlier. A preliminary search of PROSPERO, MEDLINE, the Cochrane Database of Systematic Reviews and the *JBI Evidence Synthesis* was conducted at the time of writing a priori protocol in 2022, and no current or in-progress scoping reviews or systematic reviews on the topic were identified.

In recent years, there has been a growing body of literature originating from research conducted collaboratively with and led by Indigenous communities (Chu Yang Lin et al., 2020; Murphy et al., 2021). For instance, Kirmayer and colleagues discussed community resilience and its alignment with Indigenous values and beliefs about interconnectedness of a person with their surroundings and other people (Kirmayer et al., 2009). Collaborative research methods are encouraged to promote community resilience involving both Indigenous youth and their communities (Chu Yang Lin et al., 2020). As a secondary objective, this review explored community involvement reported in the included sources that describe protective and resilience factors for Indigenous youth mental health and well-being.

### Review question(s)

The overall aim of this scoping review was to explore the resilience and protective factors that promote mental health and wellness for Indigenous youth in Canada. Specifically, our review will answer the following questions.

1. What does the literature identify as resilience or protective factors for the mental health and well-being of Indigenous youth (ages 10–25) in Canada?

a. What is the level of community involvement reported in the included studies and how is community involvement described?

### Inclusion criteria

#### Participants

In this review, we focused on research involving Indigenous youth in Canada aged 10 to 25 years. This approach aligns with the commonly adopted definition of ’youth’ in many Indigenous communities and is consistent with current mental health literature (Malla et al., 2018; World Health Organization, 2005). We also included articles that feature participants with official mental health diagnoses, self-identified mental health concerns, psychological distress symptoms, and/or involved in mental health promotion and prevention initiatives.

#### Concept

We used the following definitions to conceptualize protective and resilience factors. Protective factors referred to influences that change, mitigate, or modify an individual’s reaction to an environmental threat that increases the likelihood of an undesirable outcome (Rutter, 1985). Resilience or resilience factors referred to personality characteristics (Friborg et al., 2005), interactions with situational and contextual elements (Masten et al., 1990), or a dynamic progression that collectively contribute to favorable mental health outcomes (Luthar et al., 2000). This scoping review included all articles that explored resilience and/or protective factors for mental health and wellness of Indigenous youth. Articles exclusively concentrating on prevalence of mental health concerns without examining resilience or protective factors were excluded. It is noteworthy that articles sometimes do not explicitly reference “resilience” or “protective” factors. As such, we considered articles that explore factors that align with the definitions for resilience or protective factors mentions above which promote positive mental health outcomes.

#### Context

We considered articles conducted within healthcare, education, or community contexts in Canada. Additionally, international articles containing relevant data about Indigenous youth in Canada were also considered for inclusion. The settings encompass urban, rural, or remote areas. Articles featuring Indigenous communities or Indigenous participants residing within settler communities were also considered for inclusion. However, articles lacking extractable Canadian data were excluded.

#### Types of sources

We considered all study designs such as quantitative, qualitative, and mixed/multi-methods study for inclusion. In addition, systematic reviews, scoping reviews, commentaries, and text and opinion papers were reviewed for their reference lists to identify original studies that met our inclusion criteria. We also included grey literature such as dissertations and reports from relevant organizations that met our inclusion criteria. Due to the resources available within our research team, we included papers written in English and excluded books. No restrictions were placed on the publication dates.

## Methods

This scoping review was conducted in accordance with the JBI methodology for scoping reviews and the manuscript was prepared in line with the Preferred Reporting Items for Systematic Reviews and Meta-Analyses extension for Scoping Reviews (PRISMA-ScR). Our *a priori* has been published elsewhere previously (Carrier et al., 2022), and it is summarized below.

### Search strategy

The search strategy aimed to locate both published and unpublished primary studies, reviews, and text and opinion papers. We developed our search strategy with a health science librarian and followed the PRESS guideline (McGowan et al., 2016). An initial limited search of PubMed was undertaken to identify articles on the topic. The text words contained in the titles and abstracts of relevant articles, and the index terms used to describe the articles were used to develop a full search strategy. The search strategy, including all identified keywords and index terms was adapted for the following databases: PubMed, EMBASE, CINAHL, PsycInfo, ERIC, Scopus. The initial search was conducted on August 26th, 2021 and was updated on February 4th 2023. The full search strategies are provided in Supplementary File 1. In addition to database searches, we also conducted targeted Google search described by Godin and colleagues (Godin et al., 2015) to further identify reports and other sources such as dissertations that met our inclusion criteria. The first step involved conducting ten separate Google searches using various keyword combinations and analyzing the top 100 results from each search to identify relevant websites. Following this, the subsequent step included manually exploring the identified relevant websites to pinpoint reports and sources that align with the specified inclusion criteria. This focused Google search approach was used to complement our database searches, thus expanding the range of information sources.

### Study/Source of evidence selection

Following the search, all identified records were collated and uploaded into the Covidence (*Covidence Systematic Review Software*, 2019) and duplicates were automatically removed. Titles and abstracts were screened by two independent reviewers (HDS, AD, EC, LC, HW) for assessment against the inclusion criteria for the review. Potentially relevant papers were retrieved in full, and their citation details were imported into the Covidence. Two independent reviewers (HDS, AD, EC, HW) read full-texts papers for a detailed assessment against the inclusion criteria. Full-text papers that did not meet the inclusion criteria were excluded, and reasons for their exclusion were documented. Any disagreements that arose between the reviewers were resolved through discussion or with a third reviewer.

### Data extraction

Data were extracted from the included papers by two independent reviewers (HDS, AD, RD, RAD, HW, JL) using a data extraction tool. We first trialed the tool with three papers to assess consistency in extraction. We extracted general characteristics of the paper (e.g., author, publication country), study characteristics (e.g., aim, design, methods, setting), information on resilience and protective factors, youth characteristics (e.g., reported Indigenous communities), information on community involvement, reported outcomes and measures. See Supplementary File 2 for a full list of items in a data extraction tool. Any disagreements that arose between the reviewers were resolved through discussion or with a third reviewer.

### Data analysis and presentation

Two independent reviewers (HDS, HW) used directed content analysis (Hsieh & Shannon, 2005) to characterize extracted narrative data. First, we first organized reported protective and/or resilience factors using the adapted ecological model (Bronfenbrenner, 1994) into the following levels: 1) individual, 2) interpersonal (e.g., relationship with family and friends), 3) institutional, 4) community, 5) system and 6) wider environmental beyond social systems. The institutional level comprised organizational features, formal and informal rules, and regulations, encompassing entities like schools and hospitals. Community level encompasses the connections and interactions among organizations, institutions, and informal networks within specific boundaries. Community level also included community strengths, connection, and participation such as volunteering activities. The broader environment beyond social systems encompassed factors that extend beyond institutional and community levels, including historical, cultural, and natural elements. Protective and resilience factors were not mutually exclusive across the five levels. Second, reported community involvement was categorized following the framework called the International Association for Public Participation (IAP2) (International Association for Public Participation, 2018). This framework organizes community involvement based on the level of decision-making power held by the community in the study or project: 1) inform, 2) consult, 3) involve, 4) collaborate, 5) empower. The participating knowledge users were subsequently grouped into categories, such as elders, youth, Indigenous community members, unspecified community members, and healthcare providers. However, due to lack of reporting, coding this information into specific categories posed challenges at times. Reviewers had to interpret the information to a certain extent, relying on what the authors had reported. Any differences in interpretation were addressed through discussion between the two reviewers. We present narrative summaries along with the tables and to address our reviews questions.

## Results

### Study inclusion

Our search strategy yielded 8,318 citations (Figure 1). After Covidence automatically removed duplicates, a total of 4,641 articles were screened based on their titles and abstracts. Among these, 331 articles proceeded to full-text evaluation, ultimately leaving 55 studies that met the criteria. In addition to this, our targeted Google search yielded 6 more citations that fulfilled our inclusion criteria, bringing the total to 61 citations included in this review. The most common reasons for exclusion were ineligible population (i.e., not youth), ineligible concept, and ineligible source being review or commentaries. Full list of reasons for exclusion can be found in Figure 1.

**Figure 1:**
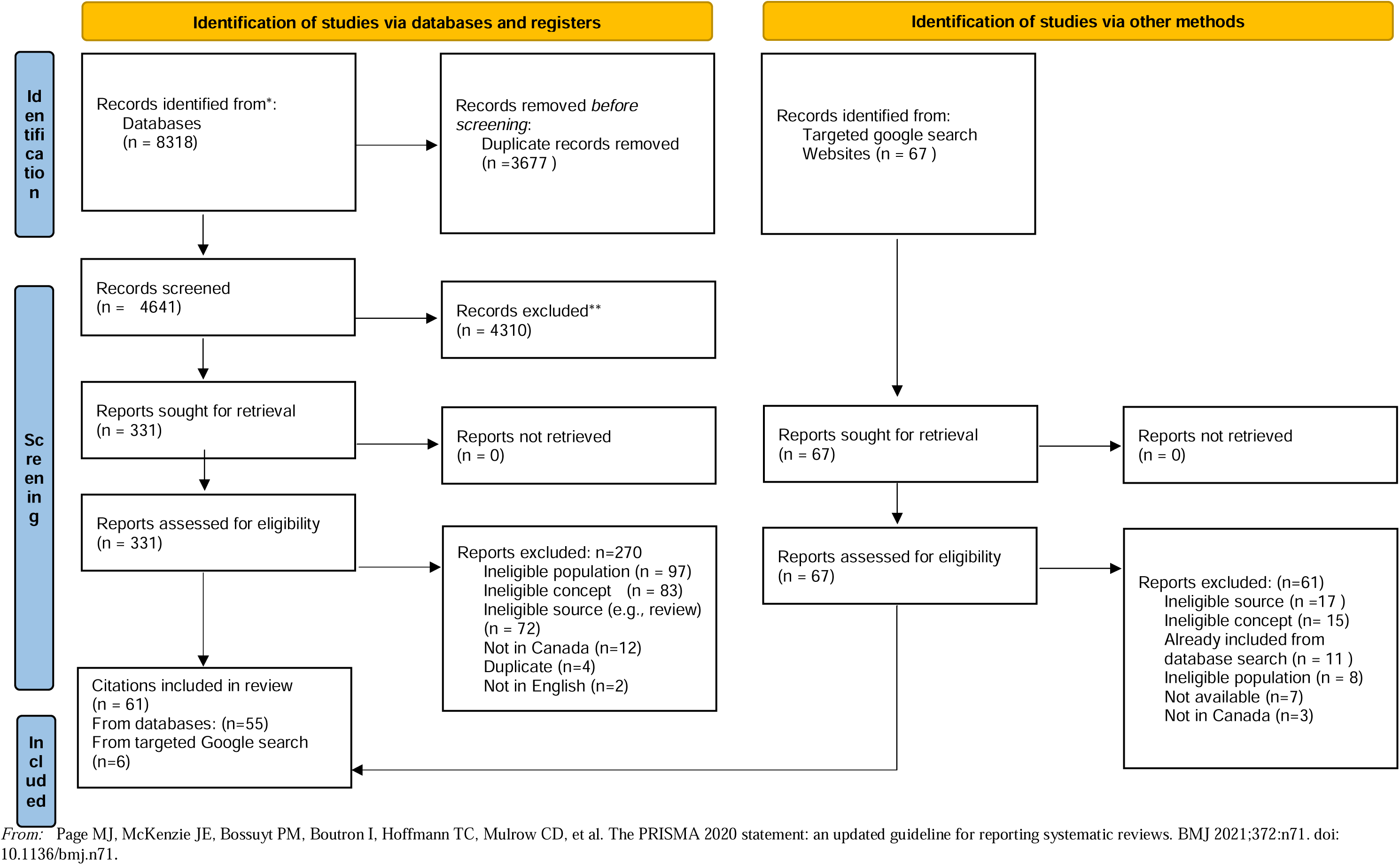
Search results and study selection and inclusion process.

### Characteristics of included studies

Individual characteristics of the included citations are organized in Supplementary File 3. As indicated in summary Table 1, 49 were peer-reviewed studies, and the remaining 12 were grey literature such as dissertations and reports. The majority were observational (n=22), following participatory action research (n=11), mixed/multi-methods (n=10), qualitative (n=9), case study (n=4), quasi-experimental (n=1), experimental (=1) and other designs such as quality improvement and program evaluation (n=3). Of the 61 citations, 31 did not report using theory, model, or framework (TMF) guiding the study or program. Twelve reported using Indigenous-specific TMFs, such as Two-Eyed Seeing, Medicine Wheel, First Nations Mental Wellness Continuum Framework, Neehithuw Child Developmental Theory, Qaujimajatuqangit (IQ) model, and Indigenous theory of planetary health. The remaining 18 reported using non-Indigenous specific TMFs, such as resilience theory, stress process model, behaviour theories, and attachment theory, competing life reinforcers model, and developmental psychopathology perspectives. There were a variety of mental health concerns for which studies or programs were examining, including substance use (n=25), general mental health and well-being (n=18), suicide (n=18), depression (n=9), anxiety (n=5), and trauma (n=3).

**Table 1.**
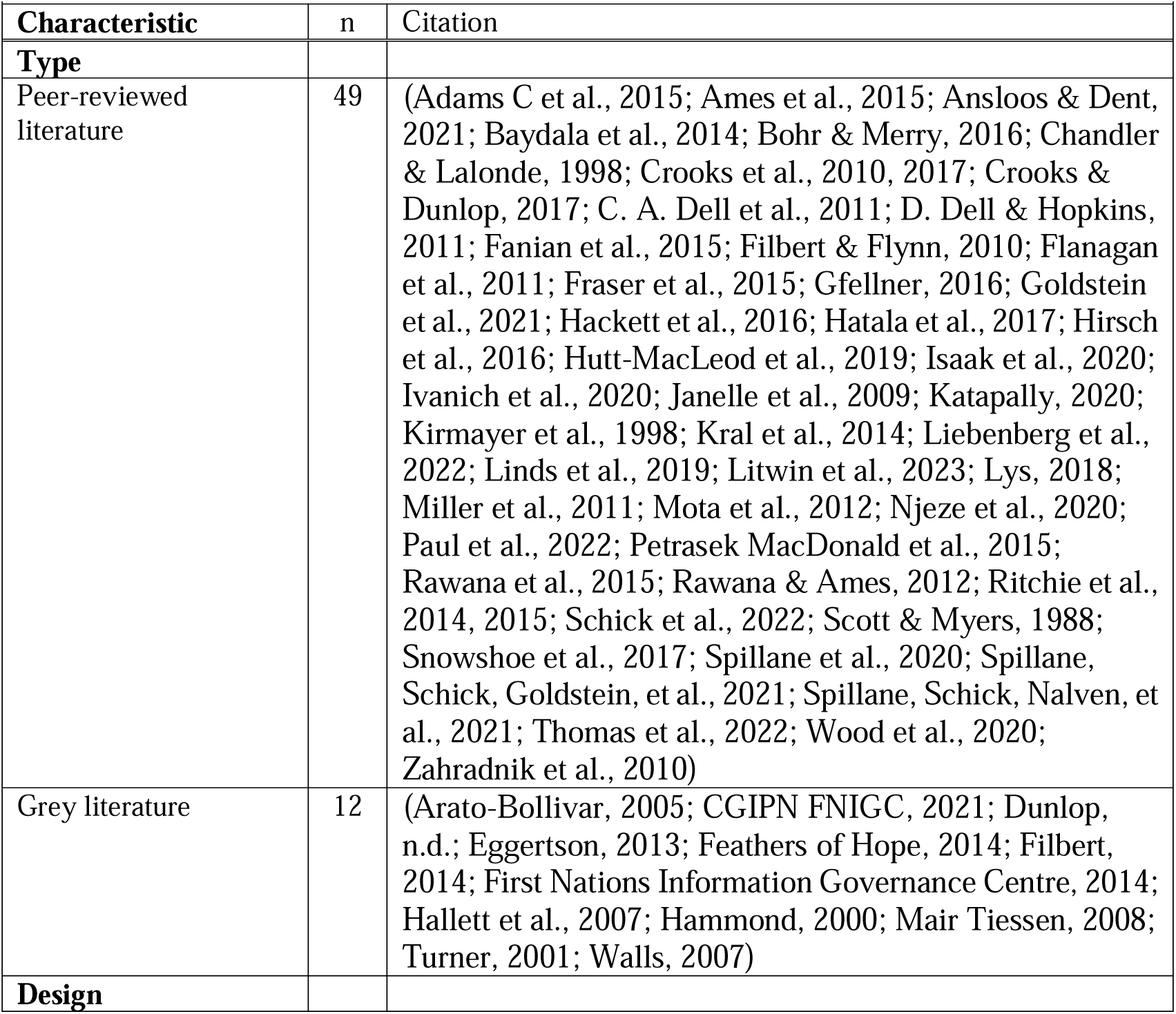
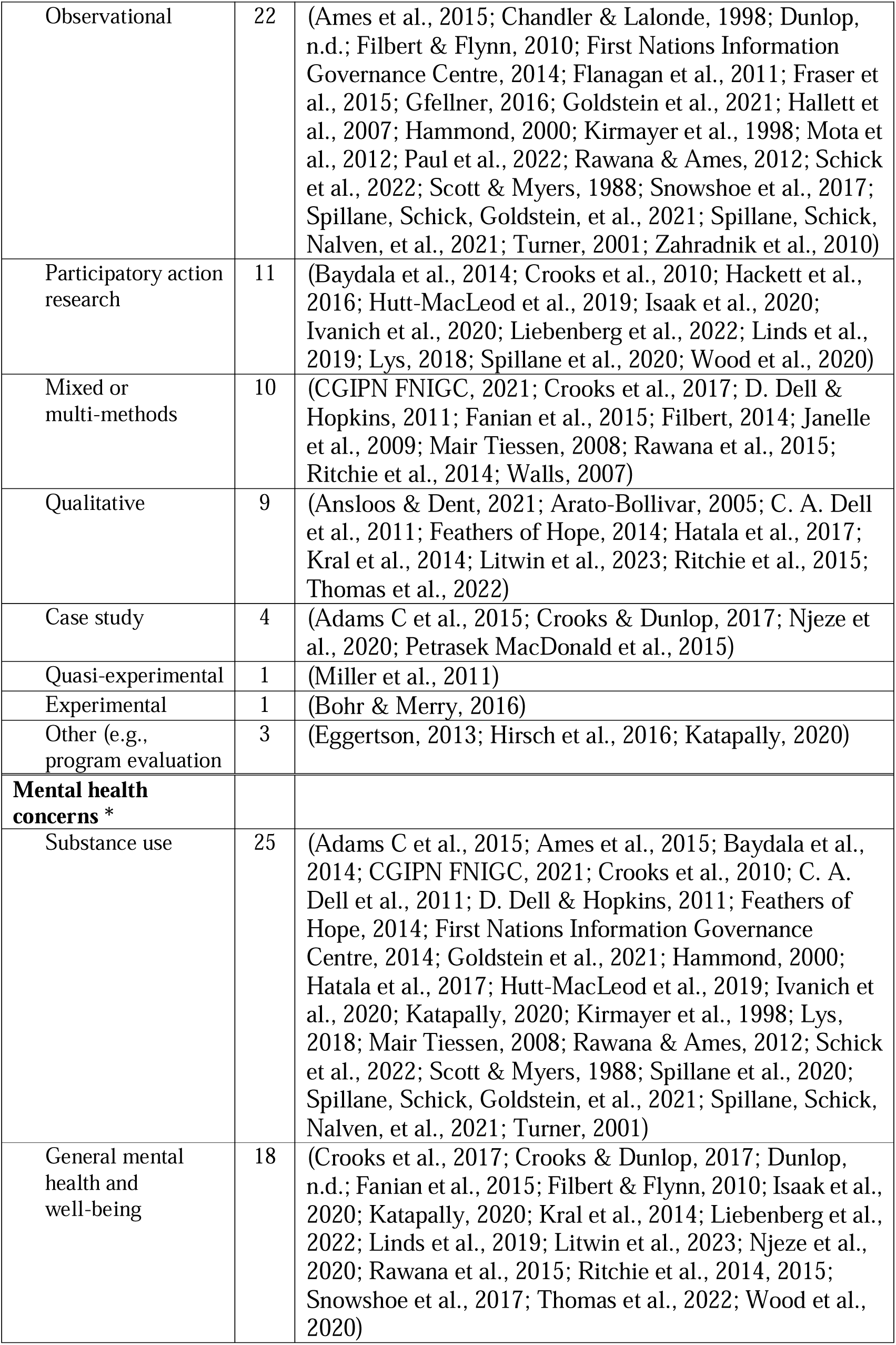
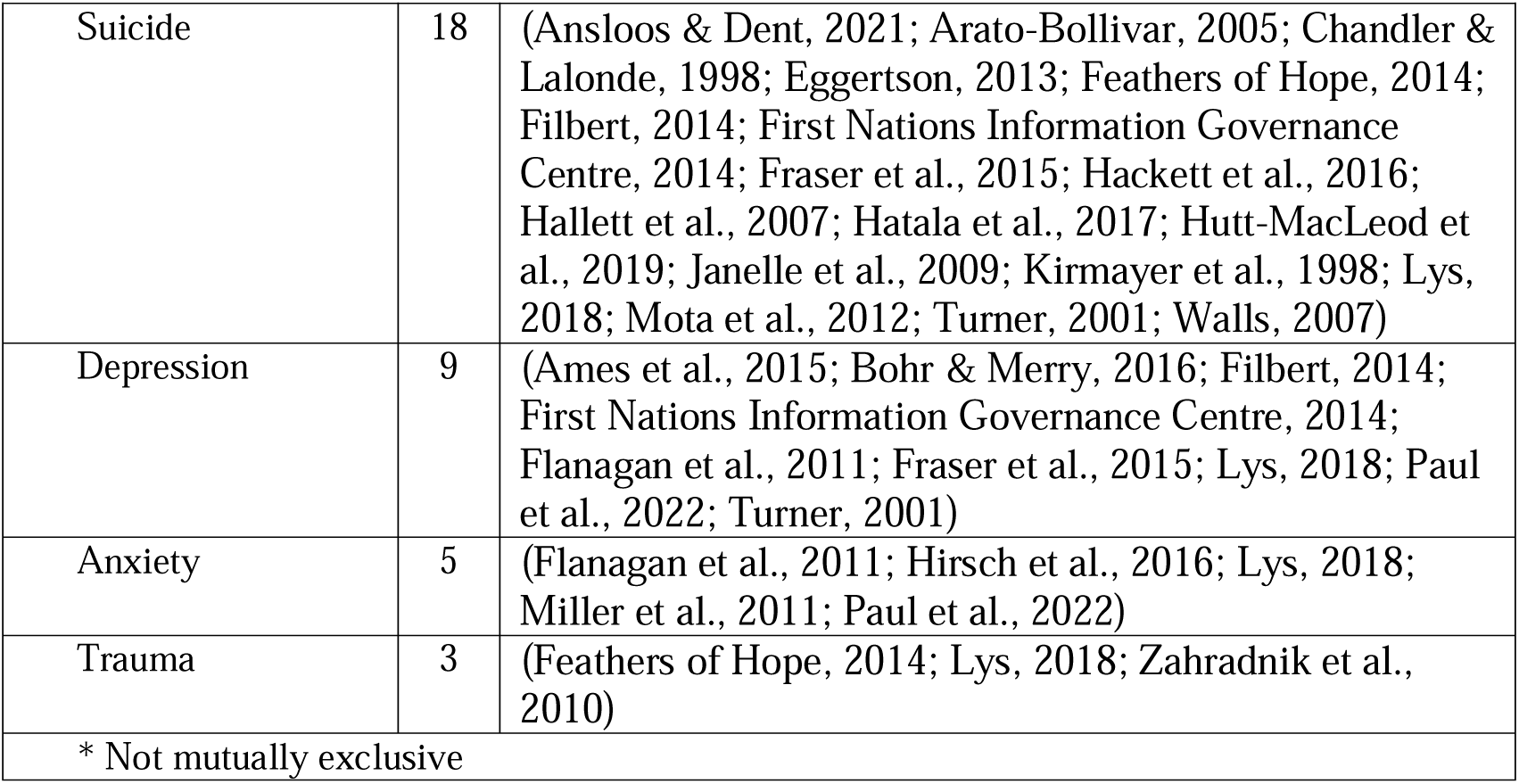
Characteristics of included literature.

Many studies took place in the Central Canada (n=23), following Prairie Provinces (n=14), Northern Canada (n=12), Atlantic region (n=10), and West coast (n=7), and two being set across Canada. Involved Indigenous communities include First Nations, not specified (n=33), Inuit (n=24), Metis (n=15), Cree (n=5), Anishinaabe - Ojibwe (n=1), Mi’kmaq (n=2), Dene (n=2), and Tłı̨chǫ(n=1).

### Review findings

#### RQ1. What does the literature identify as resilience or protective factors for the mental health and well-being of Indigenous youth (ages 10–25) in Canada?

Twenty-one citations provided definitions of protective, or resilience factors as shown in Supplementary 4. Many definitions extracted conceptualized protective and resilience factors at the individual (n=14), community (n=1), or both individual and community levels (n=4). One example definition for resilience is “An interactive process, integrating individual resources together with social and physical resources” (Wood et al., 2020, p. 392). Identified protective or resilience factors are organized into adapted socio-ecological model shown in Table 2. The three most commonly reported protective factors were found in individual level (n=52), interpersonal level (n=37), and the wider environmental level beyond social systems (n=36). Examples of individual level protective factors include self-awareness, self-confidence, self-esteem, and optimism. Examples of interpersonal level factors include relationships with family, friends and other close social networks. Examples of wider environmental level include connection to animals, plants, the Creator, ancestors and other elements in nature. Note, we characterized cultural strength and cultural identity as both individual and wider environmental level. Examples of community level factors include Indigenous community(ies) connection, community strength, and community participation, such as community volunteer activities. Of the 61 included studies, there were nine interventions or programs being implemented to promote mental health and well-being outcomes, and they include life skills training, Equine Assisted Learning program, Applied Suicide Intervention Skills training (ASIST), Substance Use Prevention Program, wilderness activity program, Smart, Positive, Active, Realistic, X-Factor thoughts (SPARX) cognitive–behavioral therapy, FRIENDS for Life cognitive behavioral program, and Outdoor Adventure Leadership Experience (OALE).

**Table 2.**
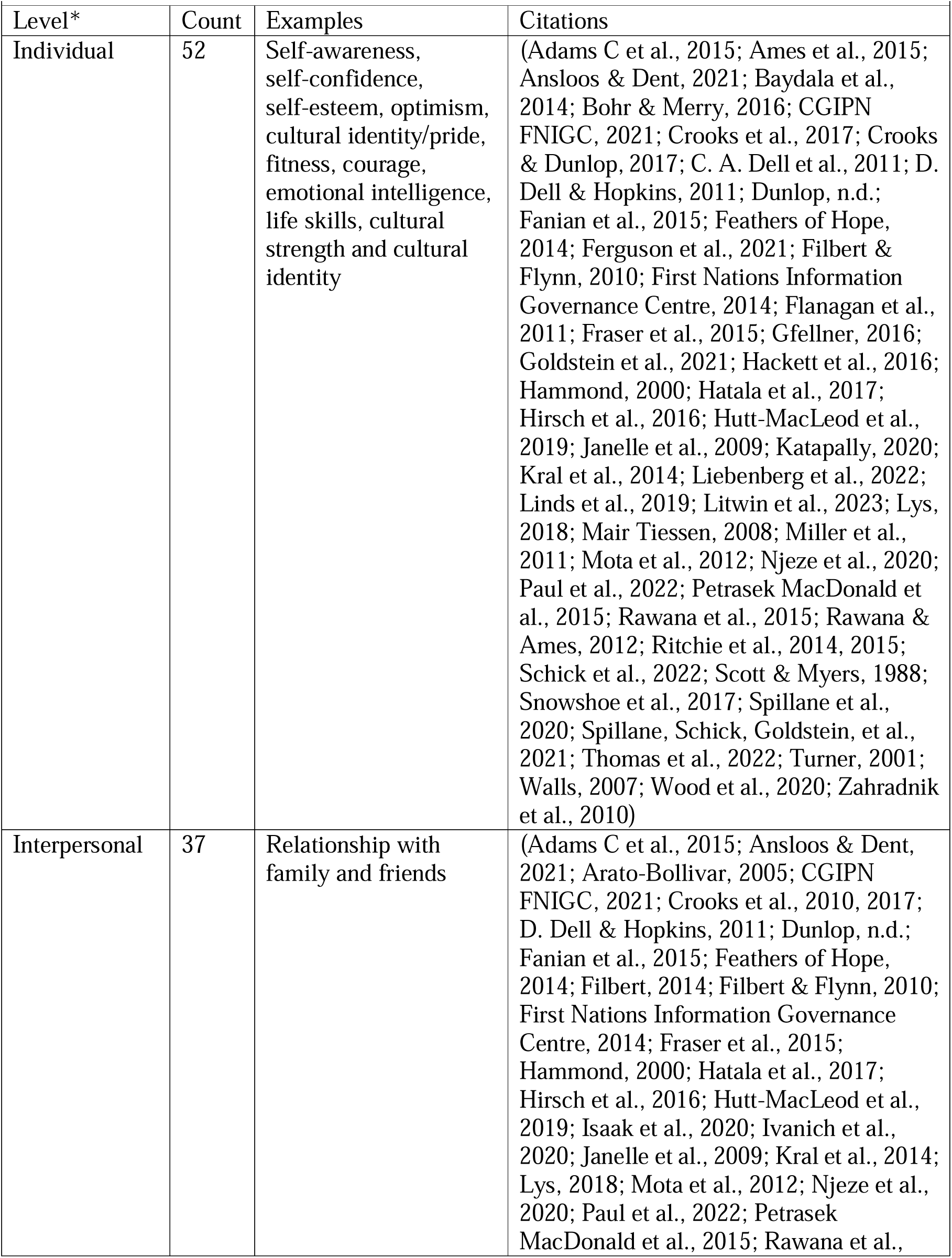
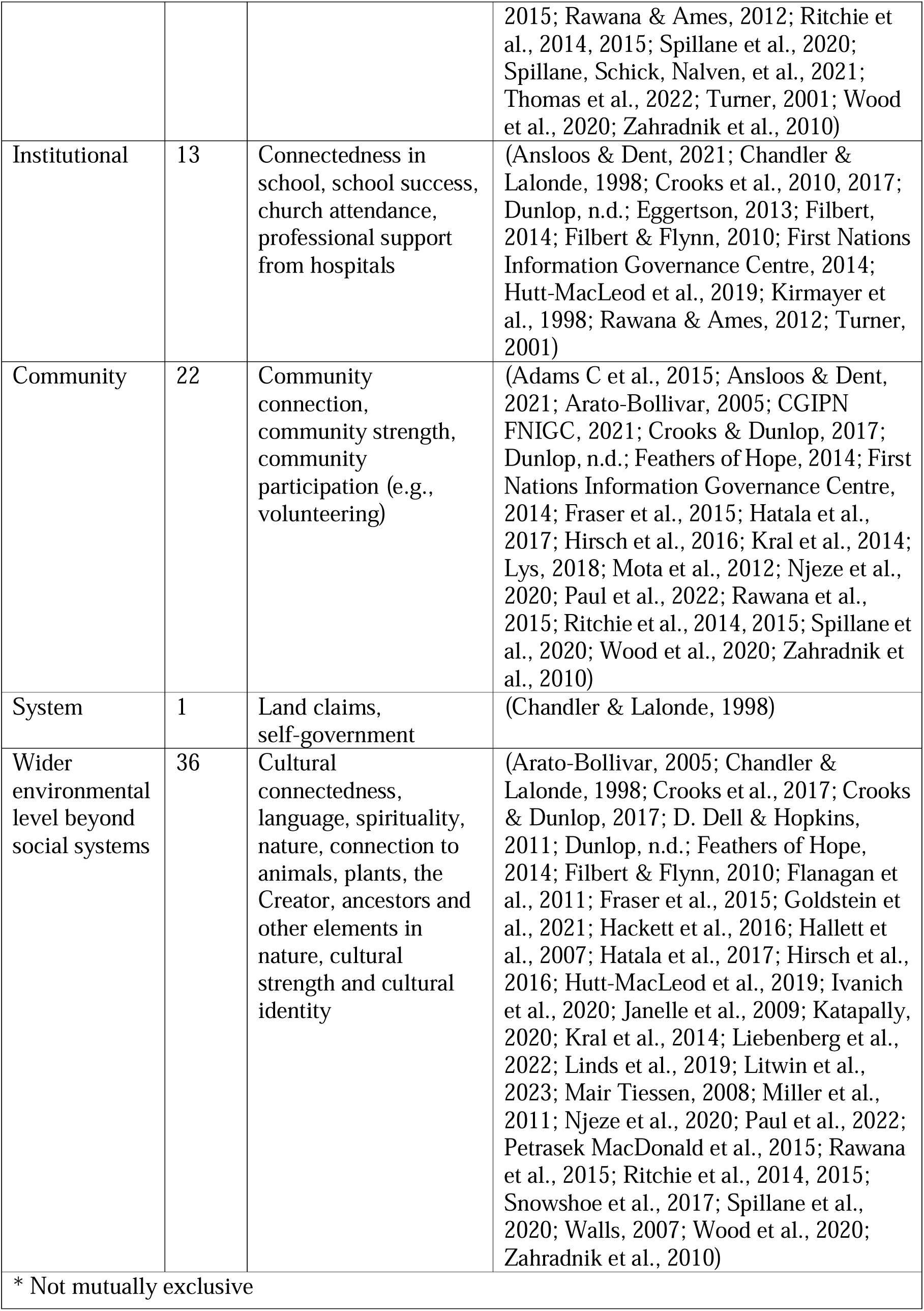
Protective and/or Resilience factors identified.

#### RQ1a. What is the level of community involvement reported in the included studies and how is community involvement described?

As shown in Figure 2, there has been a notable increase in community involvement reported in the past 10 years of publication. Forty citations provided varying detail on community involvement in their study or program, and following the IAP2 Spectrum for Public Participation, community involvement was categorized as following: inform (nlJ=lJ2); consult (nlJ=lJ7); involve (nlJ=lJ9); collaborate (nlJ=lJ17); empower (nlJ=lJ5). Notably, varying levels of involvement have been observed more frequently over the past ten years. Figure 3 presents the various levels of community involvement based on the publication year, with intervals of five years. Involved community members were non-specified community members such as friends of youth, or public citizen (n=21), youth (n=19), Indigenous community members such as community leaders and workers (n=13), Elders (n=11), family (n=8), school or program administrators including teachers (n=8), healthcare professionals (n=7) and decision makers (n=5). See Supplementary File 5 for more detailed information.

**Figure 2.**
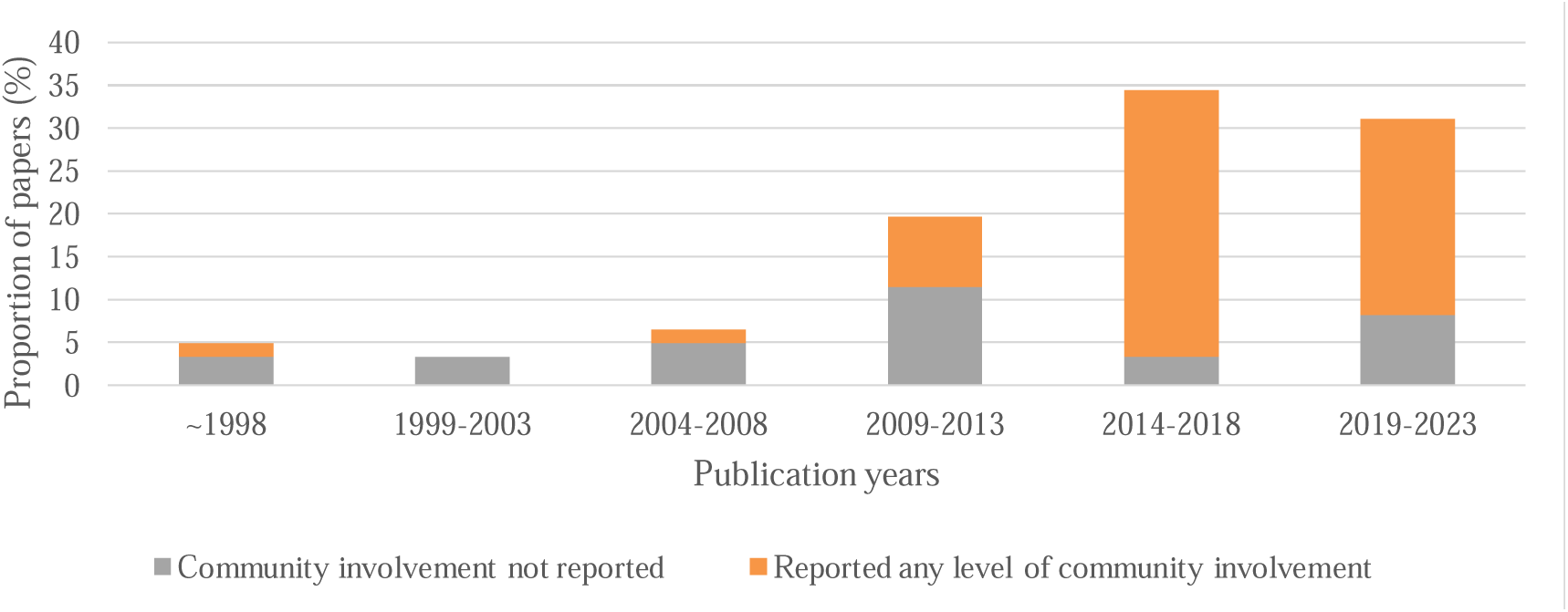
Proportion of papers that reported any level of community involvement.

**Figure 3.**
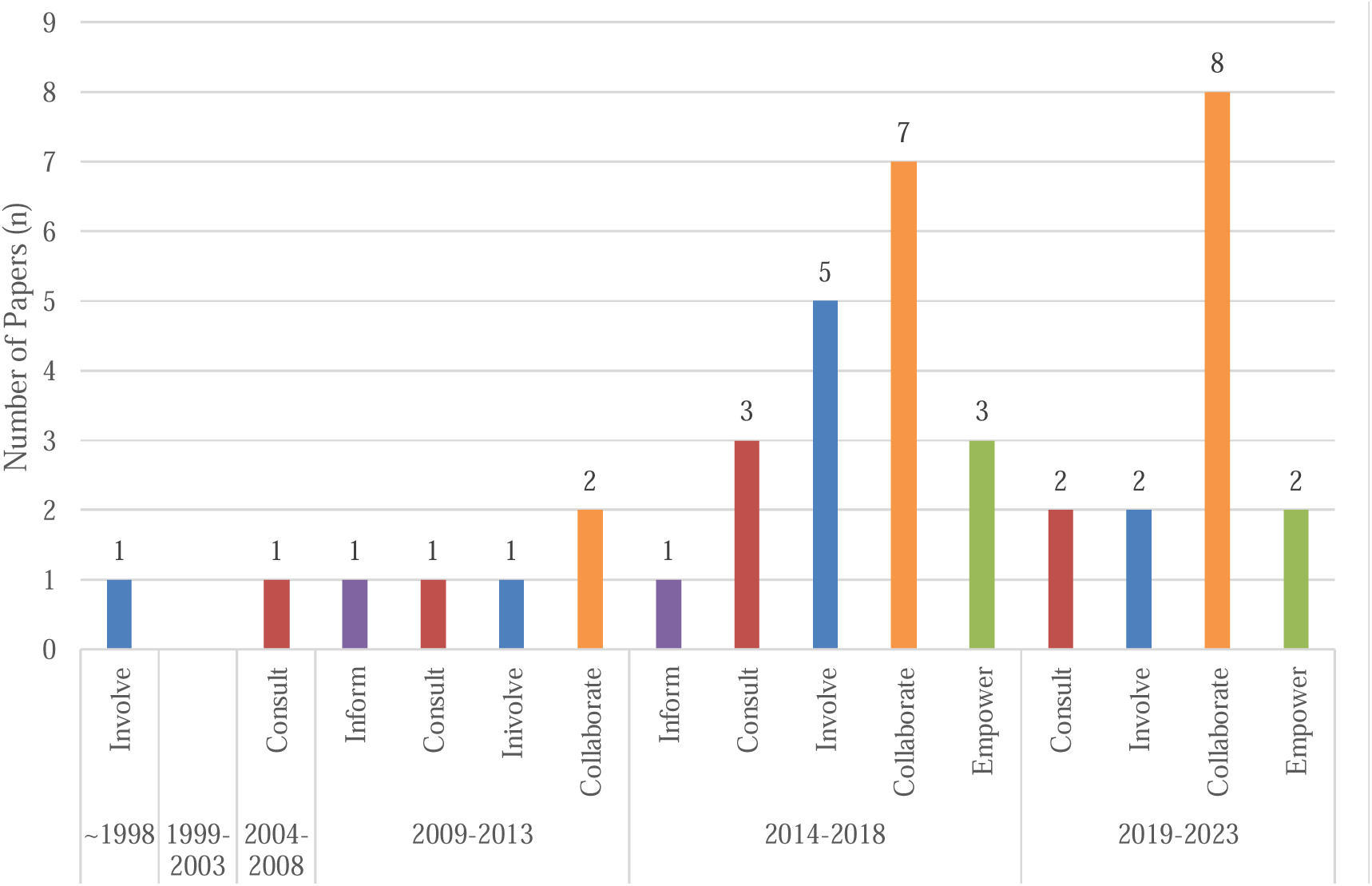
Reported levels of community involvement by publication years.

## Discussion

This scoping review identified 61 papers describing protective and/or resilience factors for diverse mental health and well-being concerns, such as substance use, suicide, and depression. Many of these papers used observational designs, followed by participatory action research approaches. Additionally, only 12 of included studies reported use of Indigenous-specific TMFs, such as Two-Eyed Seeing, to inform their projects. The study locations were primarily situated in the Central regions of Canada. The studies most commonly involved Indigenous communities including First Nations, followed by Inuit and Metis communities. Protective and resilience factors were conceptualized at the level of individual and/or community. These factors were identified across all levels, including individual, interpersonal, institutional, community, system, and wider environmental levels. There has been a marked increase in community member involvement over the past 10 years in research or programs at various stages, encompassing diverse members such as youth, Elders, Indigenous community leaders, teachers, and healthcare providers.

We found that the reported definitions of resilience and protective factors for mental health varied. While some conceptualizations of resilience were at the individual level, focusing on the ability to adapt or cope with adversity, a few papers conceptualized it at the community level, emphasizing the significance of culture and the availability of community resources. In some instances, it was understood at both levels, recognizing both individual strengths and the community’s responsibility to provide a supportive environment. Furthermore, upon examining the levels of resilience and protective factors mapped onto the socio-ecological model in the current review, multi-layered and complex nature of these factors have been revealed. Existing literature also reported varying definitions of resilience across papers, highlighting the fluid and intricate nature of the concept and the importance of adapting definitions to different Indigenous cultures and traditions (Heid et al., 2022).This intricate nature of resilience has been extensively discussed previously both nationally and internationally (Heid et al., 2022; Jongen et al., 2020; Usher et al., 2021).

It is not appropriate to use frequency counts to determine a hierarchy of resilience and protective factors for mental health and well-being (The Alliance for Child Protection in Humanitarian Action, 2021). There exist established methods for ranking protective factors, but it was beyond the scope the current review. Additionally, this review did not perform a meta-analysis to determine the overall effectiveness or causal relationships. However, it is worth noting that resilience and protective factors are most identified at both individual and wider environmental level that extends beyond social systems. Additionally, cultural strength and cultural identity have been mapped onto both the individual and the wider environmental levels. This finding aligns with current research on the significance of Indigenous youth voices and their inherent strength, illustrating how youth establish cultural connections to define and redefine their own identities and positive impact of this on their overall well-being (Barker et al., 2017; Blanchet-Cohen et al., 2023; Snowshoe et al., 2017). For example, a study in Quebec examined the various layers of meaning embedded in Indigenous youth voices (Blanchet-Cohen et al., 2023). Working closely with First Nations youth representatives and youth, the study revealed that the defining characteristics of Indigenous youth voices are strength and passion, exemplifying innate resilience in the face of adversity (Blanchet-Cohen et al., 2023). Youth described their journey of ’bringing back’ knowledge and traditions not as a way to return to the past, but as a means to draw on them in creating something new that reflects their present identity and context (Blanchet-Cohen et al., 2023). Through cultural connections, Indigenous youth understand their place in the world, find meaning in their culture, and navigate between the Western and Indigenous worlds to construct their identity being (Barker et al., 2017; Blanchet-Cohen et al., 2023; Snowshoe et al., 2017). As such, culture is crucial for identity-building and has a great connection to the promotion of mental health and well-being.

We found different levels of community involvement reported in 40 papers and ‘collaborate’ was the level most frequently inferenced based on the reported details in each paper. We also observed that before 2014, we did not find community involvement reaching the level of empowerment, which involves giving decision-making authority to Indigenous peoples rather than sharing it with researchers. This is an area in need of further exploration. While non-reporting does not necessarily imply non-engagement, it is also important to recognize that reporting is not solely the responsibility of the authors but also of the editorial boards of academic journals. A previous review report examined community involvement in 211 Indigenous research papers on all topics from the Atlantic region in Canada and found that the reported community engagement remained relatively low and steady between 2001 and 2020 but observed increased interest in requesting Indigenous ethics approvals (White et al., 2021). Additionally, mental health was one of the most frequently studied topics and found that nearly half of identified papers reported community involvement (White et al., 2021). This is similar to the current review findings, and we observed that the level of detail varied across included papers. The literature on Indigenous mental health and resiliency still exhibits a noticeable gap in reporting related to community involvement, which make the synthesis of evidence more challenging. Evaluation of collaborative approaches to research and the impact of such collaboration remains a gap in the current literature, not limited to Indigenous research (Boland et al., 2020). To facilitate clear communication and the accumulation of evidence for evaluating the impact of community involvement and to advance this field, there is a need for improved reporting and reporting guidelines, such as CONSIDER (Huria et al., 2019), are available to support researchers.

The review initially started as a class project co-led by students (HDS, LC), which eventually evolved into a project with a small grant to support it. The authors’ positionality statements are available in Supplementary File 6, providing brief self-location statements that include acknowledgments of our ancestral heritage and place-based positionality. This review was co-led by HDS and LC, and our team expanded during the process Our review adopted a collaborative and relational approach to understanding Indigenous knowledge. Recognizing some team members’ limited experience in this area, including myself, we emphasized close collaboration with those possessing Indigenous knowledge. This process reflected our commitment to humility, continuous learning, and valuing diverse perspectives. We view the review as a collective effort that honors the relationships among Indigenous, Two-Spirit, and non-Indigenous healthcare providers and researchers, highlighting our team’s foundational strength in collaboration.

Although our review team includes Indigenous nurse research trainees, we recognize the missed opportunity for community engagement in this review. The lack of community involvement in this review is a major constraint that could influence the interpretation of findings. We have highlighted how resilience and protective factors have been described in both academic and grey literature, examined the level of community involvement identified in the included sources, and identified gaps in the literature including need for better reporting. There is a future opportunity for the findings to serve as a springboard for conversations with Indigenous communities in Canada, allowing the concept of resilience and protective factors to be refined and validated. Community involvement was not initially included in the protocol, but recognizing its significance, a secondary review to incorporate community engagement to validate and potentially rank the identified factors may be valuable. Second, we would like to acknowledge that many of the frameworks we used to map our findings are of Western origin. The papers included in this review were conducted in Canada, with most of them utilizing Western methodologies to generate new knowledge. Therefore, the mapping exercise using Indigenous-specific TMFs was limited in this review, in addition to the lack of detailed reporting in the identified sources. These are important considerations when interpreting and utilizing our review findings.

## Conclusions

The papers examined here show that resilience is complex and varied in concept, and the identified heterogeneity in designs and methods confirms our appropriate choice of a scoping review methodology. Sixty-one papers included in this review discussed protective and resilience factors related to various mental health issues and well-being concerns, such as substance use, suicide, and depression. Many of these papers used observational methodologies, followed by participatory action research. Moreover, only a small number of the included studies mentioned the use of Indigenous-specific TMFs, such as Two-Eyed Seeing, to guide their projects. The majority of the study sites were located in the Central regions of Canada, primarily involving First Nations communities. Papers conceptualized protective and resilience factors at individual and/or community levels, and we mapped the identified factors across individual, interpersonal, institutional, community, system, or wider environmental level. Notably in the last decade, a range of community members, such as youth, Elders, Indigenous community leaders, and educators, participated in various research projects or programs at different levels. The causal pathways of resilience factors for mental health and well-being remain unknown in this review as meta-analysis was not the objective. However, we have highlighted the significance of Indigenous youth’s voice, one channel for demonstrating inherent strength, and their connection to culture, as well as several gaps in the literature, including the lack of reported details on community involvement. The need for resilience in the face of ongoing colonization of Indigenous Peoples persists due to systemic barriers. To address this issue, it imperative to mitigate these systemic barriers and meanwhile, examination of the protective and resilience factors highlighted in available literature offers direction for moving forward with a strengths-based focus.

## Supporting information

PRISMA-ScR

## Data Availability

All data produced in the present work are contained in the manuscript

## Acknowledgments

HDS and LC conceptualized and designed this review including the a priori protocol following the JBI methodology. This is a student-led project and HDS and LC are project leaders. HDS and LC obtained a grant to support this project and HDS managed the team. Health sciences librarians, Melissa Rothfus and Kendell Fitzgerald developed full search strategies for this review. HDS, LC, AD, EC participated in screening title and abstracts. HDS, AD, ED, HW participated in screening full texts. HDS, AD, HW, RAD, RD, JL participated in data extraction. HDS, LC, HW, RAD participated in data analysis planning. HDS and HW participated in analyzing extracted narrative data, and HDS, LC, HW, RAD participated in interpretation of results. HDS and RD developed figures and tables for data presentation. HDS drafted initial manuscript. All authors reviewed the report and gave critical feedback to the final manuscript. JAC is a supervisor of the review.

## References

Adams C, Arratoon C, Boucher J, Cartier G, Chalmers D, Dell CA, Dell D, Dryka D, Duncan R, Dunn K, Hopkins C, Longclaws L, MacKinnon T, Sauve E, Spence S, & Wuttunee M. (2015). The Helping Horse: How Equine Assisted Learning Contributes to the Wellbeing of First Nations Youth in Treatment for Volatile Substance Misuse. Hum Anim Interact Bull, 1(1), 52–75.

Ames, M. E., Rawana, J. S., Gentile, P., & Morgan, A. S. (2015). The Protective Role of Optimism and Self-esteem on Depressive Symptom Pathways Among Canadian Aboriginal Youth. Journal of Youth and Adolescence, 44(1), 142–154. 10.1007/s10964-013-0016-4

Ansloos, J., & Dent, E. (2021). “Our spirit is like a fire”: Conceptualizing intersections of mental health, wellness, and spirituality with Indigenous youth leaders across Canada | Journal of Indigenous Social Development. https://journalhosting.ucalgary.ca/index.php/jisd/article/view/72562

Arato-Bollivar, J. (2005). In their own words exploring survival factors in suicidal aboriginal youth: A critical incident study (2005-99015-002; Issues 2-A) [ProQuest Information & Learning]. http://ezproxy.library.dal.ca/login?url=https://search.ebscohost.com/login.aspx?direct=true&db=psyh&AN=2005-99015-002&site=ehost-live

Barker, B., Goodman, A., & DeBeck, K. (2017). Reclaiming Indigenous identities: Culture as strength against suicide among Indigenous youth in Canada. Canadian Journal of Public Health, 108(2), e208–e210. 10.17269/CJPH.108.5754

Baydala, L., Fletcher, F., Worrell, S., Kajner, T., Letendre, S., Letendre, L., & Rasmussen, C. (2014). Partnership, knowledge translation, and substance abuse prevention with a first nations community. *Progress in Community Health Partnerships: Research*, Education, and Action, 8(2), 145–155. 10.1353/cpr.2014.0030

Blanchet-Cohen, N., Picard, V., Robert-Careau, F., & Gray-Lehoux, C. (2023). ‘We are slowly reclaiming for ourselves’: The generative possibilities of Indigenous youth voices. Journal of Youth Studies, 1–17. 10.1080/13676261.2023.2248916

Bohr, Y., & Merry, S. (2016). Asynchronous etherapy for aboriginal communities: Experiences from Nunavut. Journal of the American Academy of Child and Adolescent Psychiatry, 55(10), S33. 10.1016/j.jaac.2016.07.561

Boland, L., Kothari, A., McCutcheon, C., Graham, I., & Integrated Knowledge Translation Research Network. (2020). Building an integrated knowledge translation (IKT) evidence base: Colloquium proceedings and research direction. Health Research Policy and Systems, 18, 1–7.

Bronfenbrenner, U. (1994). Ecological models of human development. In The International Encyclopedia of Education (2nd ed., Vol. 3). Oxford: Elsevier.

Canada, G. of C. I. S. (2019, December 11). Suicide prevention in Indigenous communities [Resource list]. https://www.sac-isc.gc.ca/eng/1576089685593/1576089741803

Carrier, L., Shin, H. D., Rothfus, M. A., & Curran, J. A. (2022). Protective and resilience factors to promote mental health among Indigenous youth in Canada: A scoping review protocol. BMJ Open, 12(1), e049285. 10.1136/bmjopen-2021-049285

CGIPN FNIGC. (2021). First Nations Youth Smoking: Factors Associated with Resilience. First Nations Information Governance Centre.

Chandler, M. J., & Lalonde, C. (1998). Cultural Continuity as a Hedge against Suicide in Canada’s First Nations. Transcultural Psychiatry, 35(2), 191–219. 10.1177/136346159803500202

Chu Yang Lin, Adalberto Loyola-Sanchez, Elaine Boyling, & Cheryl Barnabe. (2020). Community engagement approaches for Indigenous health research: Recommendations based on an integrative review. BMJ Open, 10(11), e039736. 10.1136/bmjopen-2020-039736

Covidence systematic review software. (2019). [Veritas Health Innovation]. www.covidence.org

Crooks, C. V., Chiodo, D., Thomas, D., & Hughes, R. (2010). Strengths-based Programming for First Nations Youth in Schools: Building Engagement Through Healthy Relationships and Leadership Skills. International Journal of Mental Health and Addiction, 8(2), 160–173. 10.1007/s11469-009-9242-0

Crooks, C. V., & Dunlop, C. (2017). Mental health promotion with Aboriginal youth: Lessons learned from the Uniting Our Nations program. In School mental health services for adolescents. (2017-28298-015; pp. 306–328). Oxford University Press. http://ezproxy.library.dal.ca/login?url=https://search.ebscohost.com/login.aspx?direct=true&db=psyh&AN=2017-28298-015&site=ehost-live

Crooks, C. V., Exner-Cortens, D., Burm, S., Lapointe, A., & Chiodo, D. (2017). Two Years of Relationship-Focused Mentoring for First Nations, Métis, and Inuit Adolescents: Promoting Positive Mental Health. Journal of Primary Prevention, 38(1–2), 87–104. 10.1007/s10935-016-0457-0

Dell, C. A., Chalmers, D., Bresette, N., Swain, S., Rankin, D., & Hopkins, C. (2011). A healing space: The experiences of first nations and Inuit youth with equine-assisted learning (EAL). Child & Youth Care Forum, 40(4), 319–336. 10.1007/s10566-011-9140-z

Dell, D., & Hopkins, C. (2011). Residential volatile substance misuse treatment for indigenous youth in Canada. Substance Use & Misuse, 46 *Suppl 1*((Dell D.) National Youth Solvent Abuse Committee, Saskatoon, Saskatchewan, Canada.(Hopkins C.)), 107–113. 10.3109/10826084.2011.580225

Dunlop, C. I. (n.d.). Bullying Experiences Among First Nations Youth: Identifying Effects on Mental Health and Potential Protective Factors.

Eggertson, L. (2013). Suicide prevention training saves lives in Nunavut. CMAJL: Canadian Medical Association Journal = Journal de l’Association Medicale Canadienne, 185(15), 1306–1307. 10.1503/cmaj.109-4595

Fanian, S., Young, S. K., Mantla, M., Daniels, A., & Chatwood, S. (2015). Evaluation of the Kóts’iìhtła (“we light the fire”) project: Building resiliency and connections through strengths-based creative arts programming for indigenous youth. International Journal of Circumpolar Health, 74. 10.3402/ijch.v74.27672

Feathers of Hope. (2014). Feathers of Hope: A First Nations Youth Action Plan. https://cwrp.ca/sites/default/files/publications/en/Feathers_of_Hope.pdf

Ferguson, L. J., Girolami, T., Thorstad, R., Rodgers, C. D., & Humbert, M. L. (2021). “That’s what the program is all about… building relationships”: Exploring experiences in an urban offering of the indigenous youth mentorship program in Canada. International Journal of Environmental Research and Public Health, 18(2), 1–18. 10.3390/ijerph18020733

Filbert, K. M. (2014). Developmental assets as a predictor of resilient outcomes among aboriginal young people in out-of-home care (2014-99060-224; Issues 9-B(E)) [ProQuest Information & Learning]. http://ezproxy.library.dal.ca/login?url=https://search.ebscohost.com/login.aspx?direct=true&db=psyh&AN=2014-99060-224&site=ehost-live

Filbert, K. M., & Flynn, R. J. (2010). Developmental and cultural assets and resilient outcomes in First Nations young people in care: An initial test of an explanatory model. Children and Youth Services Review, 32(4), 560–564. 10.1016/j.childyouth.2009.12.002

First Nations Information Governance Centre. (2014). Youth Resilience and Protective Factors Associated with Suicide in First Nations Communities. https://www.afnigc.ca/main/includes/media/pdf/digital%20reports/Youth%20Resilience%20FINAL%20PAPER_Mar%202014.pdf

Flanagan, T., Iarocci, G., D’Arrisso, A., Mandour, T., Tootoosis, C., Robinson, S., & Burack, J. A. (2011). Reduced ratings of physical and relational aggression for youths with a strong cultural identity: Evidence from the Naskapi people. Journal of Adolescent Health, 49(2), 155–159. 10.1016/j.jadohealth.2010.11.245

Fraser, S. L., Geoffroy, D., Chachamovich, E., & Kirmayer, L. J. (2015). Changing rates of suicide ideation and attempts among Inuit youth: A gender-based analysis of risk and protective factors. Suicide & Life-Threatening Behavior, 45(2), 141–156. PubMed. 10.1111/sltb.12122

Friborg, O., Barlaug, D., Martinussen, M., Rosenvinge, J. H., & Hjemdal, O. (2005). Resilience in relation to personality and intelligence. International Journal of Methods in Psychiatric Research, 14(1), 29–42. 10.1002/mpr.15

Gfellner, B. M. (2016). Ego strengths, racial/ethnic identity, and well-being among North American Indian/First Nations adolescents. American Indian and Alaska Native Mental Health Research, 23(3), 87–116. 10.5820/aian.2303.2016.87

Godin, K., Stapleton, J., Kirkpatrick, S. I., Hanning, R. M., & Leatherdale, S. T. (2015). Applying systematic review search methods to the grey literature: A case study examining guidelines for school-based breakfast programs in Canada. Systematic Reviews, 4(1), 138. 10.1186/s13643-015-0125-0

Goldstein, S. C., Schick, M. R., Nalven, T., & Spillane, N. S. (2021). Valuing cultural activities moderating the association between alcohol expectancies and alcohol use among first nation adolescents. Journal of Studies on Alcohol and Drugs, 82(1), 112–120. 10.15288/jsad.2021.82.112

Goulet, L., Linds, W., Episkenew, J.-A., & Schmidt, K. (2011). Creating a Space for Decolonization: Health through Theatre with Indigenous Youth. Native Studies Review, 20(1), Article 1.

Hackett, C., Furgal, C., Angnatok, D., Sheldon, T., Karpik, S., Baikie, D., Pamak, C., & Bell, T. (2016). Going Off, Growing Strong: Building Resilience of Indigenous Youth. Canadian Journal of Community Mental Health, 35(2), 79. 10.7870/cjcmh-2016-028

Hallett, D., Chandler, M. J., & Lalonde, C. E. (2007). Aboriginal language knowledge and youth suicide. Cognitive Development, 22(3), 392–399. 10.1016/j.cogdev.2007.02.001

Hammond, W. A. (2000). Canadian native adolescent solvent abuse and attachment theory (2000-95022-318; Issues 5-B) [ProQuest Information & Learning]. http://ezproxy.library.dal.ca/login?url=https://search.ebscohost.com/login.aspx?direct=true&db=psyh&AN=2000-95022-318&site=ehost-live

Hatala, A. R., Pearl, T., Bird-Naytowhow, K., Judge, A., Sjoblom, E., & Liebenberg, L. (2017). “i Have Strong Hopes for the Future”: Time Orientations and Resilience among Canadian Indigenous Youth. Qualitative Health Research, 27(9), 1330–1344. 10.1177/1049732317712489

Heid, O., Khalid, M., Smith, H., Kim, K., Smith, S., Wekerle, C., Six Nations Youth Mental Wellness Committee, Bomberry, T., Hill, L. D., & General, D. A. (2022). Indigenous youth and resilience in Canada and the USA: A scoping review. Adversity and Resilience Science, 3(2), 113–147.

Hirsch, R., Furgal, C., Hackett, C., Sheldon, T., Bell, T., Angnatok, D., Winters, K., & Pamak, C. (2016). Going off; Growing strong: A program to enhance individual youth and community resilience in the face of change in Nain, Nunatsiavut. Etudes Inuit Studies, 40(1), 63–84. 10.7202/1040145ar

Hsieh, H.-F., & Shannon, S. E. (2005). Three Approaches to Qualitative Content Analysis. Qualitative Health Research, 15(9), 1277–1288. 10.1177/1049732305276687

Huria, T., Palmer, S. C., Pitama, S., Beckert, L., Lacey, C., Ewen, S., & Smith, L. T. (2019). Consolidated criteria for strengthening reporting of health research involving indigenous peoples: The CONSIDER statement. BMC Medical Research Methodology, 19(1), 173. 10.1186/s12874-019-0815-8

Hutt-MacLeod, D., Rudderham, H., Sylliboy, A., Sylliboy-Denny, M., Liebenberg, L., Denny, J. F., Gould, M. R., Gould, N., Nossal, M., Iyer, S. N., Malla, A., & Boksa, P. (2019). Eskasoni First Nation’s transformation of youth mental healthcare: Partnership between a Mi’kmaq community and the ACCESS Open Minds research project in implementing innovative practice and service evaluation. Early Intervention in Psychiatry, 13(S1), 42–47. 10.1111/eip.12817

International Association for Public Participation (IAP2)). (2018). Core Values, Ethics, Spectrum – The 3 Pillars of Public Participation—International Association for Public Participation. Retrieved May 25, 2021, from https://cdn.ymaws.com/www.iap2.org/resource/resmgr/pillars/Spectrum_8.5x11_Print.pdf

Isaak, C. A., Mota, N., Medved, M., Katz, L. Y., Elias, B., Mignone, J., Munro, G., & Sareen, J. (2020). Conceptualizations of help-seeking for mental health concerns in First Nations communities in Canada: A comparison of fit with the Andersen Behavioral Model. Transcultural Psychiatry, 57(2), 346–362. 10.1177/1363461520906978

Isaak, C. A., Stewart, D. E., Mota, N. P., Munro, G., Katz, L. Y., & Sareen, J. (2015). Surviving, healing and moving forward: Journeys towards resilience among Canadian Cree adults. International Journal of Social Psychiatry, 61(8), 788–795. 10.1177/0020764015584648

Ivanich, J. D., Mousseau, A. C., Walls, M., Whitbeck, L., & Whitesell, N. R. (2020). Pathways of Adaptation: Two Case Studies with One Evidence-Based Substance Use Prevention Program Tailored for Indigenous Youth. Prevention Science, 21, 43–53. 10.1007/s11121-018-0914-5

Janelle, A., Laliberté, A., & Ottawa, U. (2009). Promoting traditions: An evaluation of a wilderness activity among first nations of Canada. Australasian Psychiatry, 17(SUPPL. 1), S108–S111. 10.1080/10398560902948605

Jongen, C. S., McCalman, J., & Bainbridge, R. G. (2020). A systematic scoping review of the resilience intervention literature for indigenous adolescents in CANZUS nations. Frontiers in Public Health, 7, 351.

Katapally, T. R. (2020). Smart indigenous youth: The smart platform policy solution for systems integration to address indigenous youth mental health. JMIR Pediatrics and Parenting, 3(2). 10.2196/21155

Kirmayer, L. J., Boothroyd, L. J., & Hodgins, S. (1998). Attempted suicide among Inuit youth: Psychosocial correlates and implications for prevention. Canadian Journal of Psychiatry, 43(8), 816–822. 10.1177/070674379804300806

Kirmayer, L. J., Dandeneau, S., Marshall, E., Phillips, M. K., & Williamson, K. J. (2011). Rethinking Resilience from Indigenous Perspectives. The Canadian Journal of Psychiatry, 56(2), 84–91. 10.1177/070674371105600203

Kirmayer, L. J., MD, FRCPC, Sehdev, M., Whitley, R., PhD, Dandeneau, S. F., PhD, & Isaac, C. (2009). Community Resilience: Models, Metaphors and Measures. Journal of Aboriginal Health, 5(1), 62–117. Canadian Business & Current Affairs Database; Ethnic NewsWatch.

Kral, M. J., Salusky, I., Inuksuk, P., Angutimarik, L., & Tulugardjuk, N. (2014). Tunngajuq: Stress and resilience among Inuit youth in Nunavut, Canada. Transcultural Psychiatry, 51(5), 673–692. 10.1177/1363461514533001

Liebenberg, L., Reich, J., Denny, J. F., Gould, M. R., & Hutt-MacLeod, D. (2022). Two-eyed Seeing for youth wellness: Promoting positive outcomes with interwoven resilience resources. Transcultural Psychiatry, (Liebenberg L.) Faculty of Graduate Studies, Dalhousie University, *Halifax, NS, Canada*(Reich J.*) Dalhousie University, Halifax, NS, Canada(Denny J.F.; Gould M.R.; Hutt-MacLeod D.) Eskasoni Mental Health Services, Eskasoni, NS, Can*. 10.1177/13634615221111025

Linds, W., Sjollema, S., Victor, J., Eninew, L., & Goulet, L. (2019). Widening the Angle: Film as Alternative Pedagogy for Wellness in Indigenous Youth. International Journal of Education & the Arts, 21(1), 1–29.

Litwin, L., Hankey, J., Lucassen, M., Shepherd, M., Singoorie, C., & Bohr, Y. (2023). Reflections on SPARX, a self-administered e-intervention for depression, for Inuit youth in Nunavut. Journal of Rural Mental Health, 47(1), 41–50. 10.1037/rmh0000218

Luthar, S. S., Cicchetti, D., & Becker, B. (2000). The Construct of Resilience: A Critical Evaluation and Guidelines for Future Work. Child Development, 71(3), 543–562. 10.1111/1467-8624.00164

Lys, C. (2018). Exploring coping strategies and mental health support systems among female youth in the Northwest Territories using body mapping. International Journal of Circumpolar Health, 77(1), 1466604. 10.1080/22423982.2018.1466604

MacDonald, J. P., Ford, J. D., Willox, A. C., & Ross, N. A. (2013). A review of protective factors and causal mechanisms that enhance the mental health of Indigenous Circumpolar youth. International Journal of Circumpolar Health, 72(1), 21775. 10.3402/ijch.v72i0.21775

Mair Tiessen, M. S. (2008). Collective control, cultural identity, and the psychological well-being of northern Manitoba Cree youth (2008-99220-046; Issues 5-B) [ProQuest Information & Learning]. http://ezproxy.library.dal.ca/login?url=https://search.ebscohost.com/login.aspx?direct=true&db=psyh&AN=2008-99220-046&site=ehost-live

Malla, A., Shah, J., Iyer, S., Boksa, P., Joober, R., Andersson, N., Lal, S., & Fuhrer, R. (2018). Youth Mental Health Should Be a Top Priority for Health Care in Canada. The Canadian Journal of Psychiatry, 63(4), 216–222. 10.1177/0706743718758968

Masten, A. S., Best, K. M., & Garmezy, N. (1990). Resilience and development: Contributions from the study of children who overcome adversity. Development and Psychopathology, 2(4), 425–444. Cambridge Core. 10.1017/S0954579400005812

McGowan, J., Sampson, M., Salzwedel, D. M., Cogo, E., Foerster, V., & Lefebvre, C. (2016). PRESS Peer Review of Electronic Search Strategies: 2015 Guideline Statement. Journal of Clinical Epidemiology, 75, 40–46. 10.1016/j.jclinepi.2016.01.021

Miller, L. D., Laye-Gindhu, A., Bennett, J. L., Liu, Y., Gold, S., March, J. S., Olson, B. F., & Waechtler, V. E. (2011). An effectiveness study of a culturally enriched school-based CBT anxiety prevention program. Journal of Clinical Child and Adolescent Psychology, 40(4), 618–629. 10.1080/15374416.2011.581619

Mota, N., Elias, B., Tefft, B., Medved, M., Munro, G., & Sareen, J. (2012). Correlates of suicidality: Investigation of a representative sample of Manitoba First Nations adolescents. American Journal of Public Health, 102(7), 1353–1361. PubMed. 10.2105/AJPH.2011.300385

Murphy, K., Branje, K., White, T., Cunsolo, A., Latimer, M., McMillan, J., Sylliboy, J. R., McKibbon, S., & Martin, D. (2021). Are we walking the talk of participatory Indigenous health research? A scoping review of the literature in Atlantic Canada. PLoS One, 16(7), e0255265.

Njeze, C., Bird-Naytowhow, K., Pearl, T., & Hatala, A. R. (2020). Intersectionality of Resilience: A Strengths-Based Case Study Approach With Indigenous Youth in an Urban Canadian Context. Qualitative Health Research, 30(13), 2001–2018. 10.1177/1049732320940702

Paul, J., McQuaid, R. J., Hopkins, C., Perri, A., Stewart, S., Matheson, K., Anisman, H., & Bombay, A. (2022). Relations between bullying and distress among youth living in First Nations communities: Assessing direct and moderating effects of culture-related variables. Transcultural Psychiatry*, (*Paul J.; Stewart S.) Department of Psychology and Neuroscience, Dalhousie University, *Halifax, Canada*(McQuaid R.J.; Matheson K.; Anisman H.*) Department of Neuroscience, Carleton University, Ottawa, Canada(*McQuaid R.J.*) Institute of Mental Health Research*. 10.1177/13634615221109359

Peters, P. A., Oliver, L. N., & Kohen, D. E. (2013). Mortality among children and youth in high-percentage First Nations identity areas, 2000-2002 and 2005-2007. Rural and Remote Health, 13(3), 2424–2424.

Petrasek MacDonald, J., Cunsolo Willox, A., Ford, J. D., Shiwak, I., & Wood, M. (2015). Protective factors for mental health and well-being in a changing climate: Perspectives from Inuit youth in Nunatsiavut, Labrador. Social Science & Medicine, 141, 133–141. 10.1016/j.socscimed.2015.07.017

Rawana, J. S., & Ames, M. E. (2012). Protective Predictors of Alcohol Use Trajectories Among Canadian Aboriginal Youth. Journal of Youth and Adolescence, 41(2), 229–243. 10.1007/s10964-011-9716-9

Rawana, J. S., Sieukaran, D. D., Nguyen, H. T., & Pitawanakwat, R. (2015). Development and evaluation of a peer mentorship program for Aboriginal university students. Canadian Journal of Education, 38(2), 1–34. 10.2307/canajeducrevucan.38.2.08

Ritchie, S. D., Wabano, M. J., Corbiere, R. G., Restoule, B. M., Russell, K. C., & Young, N. L. (2015). Connecting to the Good Life through outdoor adventure leadership experiences designed for Indigenous youth. Journal of Adventure Education and Outdoor Learning, 15(4), 350–370. 10.1080/14729679.2015.1036455

Ritchie, S. D., Wabano, M.-J., Russell, K., Enosse, L., & Young, N. L. (2014). Promoting resilience and wellbeing through an outdoor intervention designed for Aboriginal adolescents. Rural and Remote Health, 14, 2523.

Rowhani, M., & Hatala, A. (2017). A Systematic Review of Resilience Research among Indigenous Youth in Contemporary Canadian Contexts. *The International Journal of Health*, Wellness, and Society, 7, 45–58. 10.18848/2156-8960/CGP/v07i04/45-58

Rutter, M. (1985). Resilience in the face of adversity. Protective factors and resistance to psychiatric disorder. The British Journal of PsychiatryL: The Journal of Mental Science, 147, 598–611. PubMed. 10.1192/bjp.147.6.598

Schick, M. R., Todi, A. A., & Spillane, N. S. (2022). Subjective Happiness Interrupts the Association Between Alcohol Expectancies and Alcohol Consumption Among Reserve-Dwelling First Nation Adolescents. American Journal of Orthopsychiatry, 92(4), 497–504. 10.1037/ort0000607

Scott, K. A., & Myers, A. M. (1988). Impact of fitness training on native adolescents’ self-evaluations and substance use. Canadian Journal of Public Health, 79(6), 424–429.

Sikorski, C., Leatherdale, S., & Cooke, M. (2019). Tobacco, alcohol and marijuana use among Indigenous youth attending off-reserve schools in Canada: Cross-sectional results from the Canadian Student Tobacco, Alcohol and Drugs Survey. Maladies Chroniques et Blessures Au Canada, 39.

Smylie, J., Fell, D., Ohlsson, A., & Joint Working Group on First Nations, I., Inuit, and Métis Infant Mortality of the Canadian Perinatal Surveillance System. (2010). A Review of Aboriginal Infant Mortality Rates in Canada: Striking and Persistent Aboriginal/Non-Aboriginal Inequities. Canadian Journal of Public Health, 101(2), 143–148. 10.1007/BF03404361

Snowshoe, A., Crooks, C. V., Tremblay, Paul. F., & Hinson, R. E. (2017). Cultural Connectedness and Its Relation to Mental Wellness for First Nations Youth. The Journal of Primary Prevention, 38(1), 67–86. 10.1007/s10935-016-0454-3

Spillane, N. S., Kirk-Provencher, K. T., Schick, M. R., Nalven, T., Goldstein, S. C., & Kahler, C. W. (2020). Identifying competing life reinforcers for substance use in First Nation adolescents. Substance Use & Misuse, 55(6), 886–895. 10.1080/10826084.2019.1710206

Spillane, N. S., Schick, M. R., Goldstein, S. C., Nalven, T., & Kahler, C. W. (2021). The protective effects of self-compassion on alcohol-related problems among first nation adolescents. Addiction Research and Theory. 10.1080/16066359.2021.1902994

Spillane, N. S., Schick, M. R., Nalven, T., Goldstein, S. C., Kirk-Provencher, K. T., Hill, D., & Kahler, C. W. (2021). Testing the competing life reinforcers model for substance use in reserve-dwelling First Nation youth. American Journal of Orthopsychiatry, 91(4), 477–486. 10.1037/ort0000543

The Alliance for Child Protection in Humanitarian Action. (2021, May 3). Guidance | Identifying and Ranking Risk and Protective Factors: A Brief Guide | Alliance CHPA. https://alliancecpha.org/en/child-protection-online-library/guidance-identifying-and-ranking-risk-and-protective-factors-brief

Thomas, A., Bohr, Y., Hankey, J., Oskalns, M., Barnhardt, J., & Singoorie, C. (2022). How did Nunavummiut youth cope during the COVID-19 pandemic? A qualitative exploration of the resilience of Inuit youth leaders involved in the I-SPARX project. International Journal of Circumpolar Health, 81(1), 2043577. 10.1080/22423982.2022.2043577

Toombs, E., Kowatch, K. R., & Mushquash, C. J. (2016). Resilience in Canadian Indigenous youth: A scoping review. International Journal of Child and Adolescent Resilience, 4(1), 4–32.

Tousignant, M., PhD, & Sioui, N., PhD. (2009). Resilience and Aboriginal Communities in Crisis: Theory and Interventions. Journal of Aboriginal Health, 5(1), 43–61. Canadian Business & Current Affairs Database; Ethnic NewsWatch.

Turner, R. A. (2001). Risk protective factors for propensity for suicide among British Columbia First Nations adolescents using the Adolescent Health Survey (2001-95002-138; Issues 7-B) [ProQuest Information & Learning]. http://ezproxy.library.dal.ca/login?url=https://search.ebscohost.com/login.aspx?direct=true&db=psyh&AN=2001-95002-138&site=ehost-live

Ungar, M., & Theron, L. (2020). Resilience and mental health: How multisystemic processes contribute to positive outcomes. The Lancet Psychiatry, 7(5), 441–448.

Usher, K., Jackson, D., Walker, R., Durkin, J., Smallwood, R., Robinson, M., Sampson, U. N., Adams, I., Porter, C., & Marriott, R. (2021). Indigenous resilience in Australia: A scoping review using a reflective decolonizing collective dialogue. Frontiers in Public Health, 9, 630601.

Walls, M. L. (2007). A mixed methods examination of indigenous youth suicide (2007-99230-237; Issues 6-A) [ProQuest Information & Learning]. http://ezproxy.library.dal.ca/login?url=https://search.ebscohost.com/login.aspx?direct=true&db=psyh&AN=2007-99230-237&site=ehost-live

White, T., Murphy, K., Branje, K., McKibbon, S., Cunsolo, A., Latimer, M., McMillan, J., Sylliboy, J., & Martin, D. (2021). How has Indigenous Health Research changed in Atlantic Canada over two decades? A scoping review from 2001 to 2020. Social Science & Medicine, 279,113947. 10.1016/j.socscimed.2021.113947

Wood, M., Liebenberg, L., Ikeda, J., Vincent, A., & Youth participants of Spaces & Places. (2020). The Role of Educational Spaces in Supporting Inuit Youth Resilience. Child Care in Practice, 26(4), 390–415. 10.1080/13575279.2020.1765143

World Health Organization. (2005). Atlas: Child and adolescent mental health resources: Global concerns: Implications for the future. Child and Adolescent Mental Health Atlas. WHO IRIS. https://apps.who.int/iris/handle/10665/43307

Zahradnik, M., Stewart, S. H., O’Connor, R. M., Stevens, D., Ungar, M., & Wekerle, C. (2010). Resilience Moderates the Relationship Between Exposure to Violence and Posttraumatic Reexperiencing in Mi’kmaq Youth. International Journal of Mental Health and Addiction, 8(2), 408–420. 10.1007/s11469-009-9228-y

