## Supplementary File 1 for "Resilience and Protective Factors for Mental Health among Indigenous Youth in Canada: A Scoping Review"

**APPENDIX 1: Search strategy**

**PubMed**

Initial searches conducted on August 26, 2021

| **Search** | **Query** | **Records retrieved** |
| --- | --- | --- |
| #1 | ("Adolescent"[Mesh] OR "Young Adult"[Mesh] OR "Child"[Mesh]) AND "Indigenous Canadians"[Mesh] | 1,498 |
| #2 | (child*[tiab] OR youth*[tiab] OR adolescent*[tiab] OR "school age"[tiab] OR "middle school"[tiab] OR "junior high"[tiab] OR "high school"[tiab] OR teen*[tiab] OR "young adult"[tiab] OR “emerging adult”[tiab]) AND ("first nations"[tiab] OR "inuit"[tiab] OR metis[tiab] OR aboriginal*[tiab] OR indigenous[tiab] OR "first people"[tiab] OR native[tiab]) | 13,565 |
| #3 | #1 OR #2 | 14,648 |
| #4 | (Arctic[tw]) OR (circumpolar[tw]) OR (“Prince Edward Island"[tw]) OR ("Newfoundland and Labrador"[tw]) OR ("Nova Scotia*"[tw]) OR (Canadian*[tw]) OR (Canada[tw]) OR ("P.E.I."[tw]) OR ("New Brunswick"[tw]) OR ("Quebec*"[tw]) OR ("Ontario"[tw]) OR ("Manitoba*"[tw]) OR ("Saskatchewan"[tw]) OR ("Alberta*"[tw]) OR ("British Columbia*"[tw]) OR ("B.C."[tw]) OR ("Northwest Territories"[tw]) OR ("N.W.T."[tw]) OR ("Nunavut"[tw]) OR ("Yukon Territory"[tw]) | 285,973 |
| #5 | “Arctic regions”[Mesh] OR "Canada"[Mesh] OR "Nova Scotia"[Mesh] OR "Newfoundland and Labrador"[Mesh] OR "Prince Edward Island"[Mesh] OR "New Brunswick"[Mesh] OR "Quebec"[Mesh] OR "Ontario"[Mesh] OR "Manitoba"[Mesh] OR "Saskatchewan"[Mesh] OR "Alberta"[Mesh] OR "British Columbia"[Mesh] OR "Northwest Territories"[Mesh] OR "Nunavut"[Mesh] OR "Yukon Territory"[Mesh] | 174,128 |
| #6 | #4 OR #5 | 285,973 |
| #7 | (“Suicide”[Mesh]) OR (“Mental Health"[Mesh]) OR ("Mental Disorders"[Mesh]) OR ("Psychology"[Mesh:NoExp]) OR ("Psychology, Adolescent"[Mesh]) OR ("Resilience, Psychological"[Mesh]) OR ("Community Mental Health Services"[Mesh]) OR ("Culturally Competent Care"[Mesh]) OR ("Health Services"[Mesh]) OR ("Health Services Accessibility"[Mesh]) OR ("Psychological Distress"[Mesh]) OR ("Behavioral Symptoms"[Mesh]) OR ("Substance-Related Disorders"[Mesh]) | 3,713,983 |
| #8 | “mental* ill*”[tiab] OR suicid*[tiab] OR resilen*[tiab] OR adaptability[tiab] OR "protective factors"[tiab] OR "mental health"[tiab] OR coping[tiab] OR cope[tiab] OR "mental wellness"[tiab] OR "mental wellbeing" OR psycho-social[tiab] OR psychosocial[tiab] OR trauma*[tiab] OR “cultural safety” [tiab] OR “cultural competence”[tiab] OR "social support"[tiab] OR “mental toughness”[tiab] OR “community involvement”[tiab] OR “community support*”[tiab] OR “psychosis”[tiab] OR “mental disorder*”[tiab] OR “substance-related disorder*”[tiab] OR “substance abuse” [tiab] OR “substance use*”[tiab] OR “drug use*”[tiab] OR “drug abuse”[tiab] OR “drug dependence”[tiab] OR alcoholism[tiab] OR “alcohol abuse”[tiab] OR “disruptive behavior*”[tiab] OR “disruptive behaviour*”[tiab] OR "Addiction"[tiab] OR "Alcohol Use"[tiab] OR "Cannabis Use*"[tiab] OR “cannabis abuse”[tiab] OR "Drug Dependenc*"[tiab] OR "Inhalant Abuse"[tiab] OR “inhalant use*”[tiab] OR “opioid abuse”[tiab] OR "Opioid Use*"[tiab] OR "Tobacco Use*”[tiab] OR “prescription abuse”[tiab] OR “chronic stress”[tiab] OR “acute stress”[tiab] OR “adjustment disorder*”[tiab] OR “stress disorder*”[tiab] OR depression[tiab] OR “depressive symptoms”[tiab] | 1,441,683 |
| #9 | #7 OR #8 | 4,429,175 |
| #10 | #3 AND #6 AND #9 | 1,024 |

**EMBASE**

Searches conducted on August 26, 2021

| **Search** | **Query** | **Records Retrieved** |
| --- | --- | --- |
| #1 | ('adolescent'/exp OR 'young adult'/exp OR ‘child’/exp OR 'middle school student'/exp OR 'high school student'/exp) AND ('Inuit'/exp OR 'First Nation'/exp) | 1,252 |
| #2 | (child*:ti,ab OR youth*:ti,ab OR adolescent*:ti,ab OR ‘school age’:ti,ab OR ‘middle school’:ti,ab OR ‘junior high’:ti,ab OR ‘high school’:ti,ab OR teen*:ti,ab OR ‘young adult’:ti,ab OR ‘emerging adult’:ti,ab) AND (inuit:ti,ab OR metis:ti,ab OR aboriginal*:ti,ab OR indigenous:ti,ab OR ‘first people’:ti,ab OR native:ti,ab) | 16,938 |
| #3 | #1 OR #2 | 17,642 |
| **#4** | ‘Canada’/exp OR ‘arctic region’/exp | 200,995 |
| **#5** | Arctic:ti,ab,kw OR circumpolar:ti,ab,kw OR ‘Prince Edward Island’:ti,ab,kw OR ‘Newfoundland and Labrador’:ti,ab,kw OR ‘Nova Scotia*’:ti,ab,kw OR Canadian*:ti,ab,kw OR Canada:ti,ab,kw OR ‘P.E.I.’:ti,ab,kw OR ‘New Brunswick’:ti,ab,kw OR Quebec:ti,ab,kw OR Ontario:ti,ab,kw OR Manitoba*:ti,ab,kw OR Saskatchewan:ti,ab,kw OR Alberta*:ti,ab,kw OR ‘British Columbia*’:ti,ab,kw OR ‘B.C.’:ti,ab,kw OR ‘Northwest Territories’:ti,ab,kw OR ‘N.W.T.’:ti,ab,kw OR Nunavut:ti,ab,kw OR ‘Yukon Territory’:ti,ab,kw | 288,706 |
| **#6** | #4 OR #5 | 350,592 |
| **#7** | ‘Suicide’/exp OR ‘Mental Health’/exp OR ‘Mental Disease’/exp OR ‘Psychology’/de OR ‘Psychological Resilience’/exp OR 'community mental health service'/exp OR 'cultural competence'/exp OR 'health service'/exp OR 'health care access'/exp OR 'drug dependence'/exp OR 'child psychiatry'/exp OR 'disruptive behavior'/exp OR 'acute psychosis'/exp | 8,104,052 |
| **#8** | ‘mental* ill*’:ti,ab OR ‘suicide’:ti,ab OR ‘child psychology’:ti,ab OR resilen*:ti,ab OR ‘protective factors’:ti,ab OR ‘mental health’:ti,ab OR coping:ti,ab OR cope:ti,ab OR ‘mental wellness’:ti,ab OR ‘mental wellbeing’:ti,ab OR ‘psycho-social’:ti,ab OR psychosocial:ti,ab OR ‘cultural safety’:ti,ab OR ‘cultural competence’:ti,ab OR ‘social support’:ti,ab OR trauma*:ti,ab OR ‘cultur* connectedness’:ti,ab OR ‘mental toughness’:ti,ab OR adaptability:ti,ab OR ‘community involvement’:ti,ab OR ‘community support*’:ti,ab OR ‘psychosis’:ti,ab OR ‘mental disorder*’:ti,ab OR ‘substance-related disorder*’:ti,ab OR ‘substance abuse’:ti,ab OR ‘substance use*’:ti,ab OR ‘drug use*’:ti,ab OR ‘drug abuse’:ti,ab OR ‘drug dependence’:ti,ab OR alcoholism:ti,ab OR ‘alcohol abuse’:ti,ab OR ‘disruptive behavior*’:ti,ab OR ‘disruptive behaviour*’:ti,ab OR ‘Addiction’:ti,ab OR ‘Alcohol Use’:ti,ab OR ‘Cannabis Use’:ti,ab OR ‘cannabis abuse’:ti,ab OR ‘Drug Dependenc*’:ti,ab OR ‘Inhalant Abuse’:ti,ab OR ‘inhalant use*’:ti,ab OR ‘opioid abuse’:ti,ab OR ‘Opioid Use’:ti,ab OR ‘Tobacco Use Disorder’:ti,ab OR ‘prescription abuse’:ti,ab OR ‘chronic stress’:ti,ab OR ‘acute stress’:ti,ab OR ‘adjustment disorder*’:ti,ab OR ‘stress disorder*’:ti,ab OR depression:ti,ab OR ‘depressive symptoms’:ti,ab | 1,841,855 |
| **#9** | #7 OR #8 | 8,728,982 |
| **#10** | #3 AND #6 AND #9 | 1,165 |

**CINAHL**

Searches conducted on August 26, 2021

| **Search** | **Query** | **Records Retrieved** |
| --- | --- | --- |
| **#1** | TI ((child* OR youth* OR adolescent* OR “school age” OR “middle school” OR “junior high” OR “high school” OR teen* OR “young adult” OR “emerging adult”) AND (“first nations” OR “inuit” OR “metis” OR “aboriginal*” OR “indigenous” OR “first people” OR native))  OR AB ((child* OR youth* OR adolescent* OR “school age” OR “middle school” OR “junior high” OR “high school” OR teen* OR “young adult” OR “emerging adult”) AND (“first nations” OR “inuit” OR “metis” OR “aboriginal*” OR “indigenous” OR “first people” OR native)) | 5,614 |
| **#2** | (((MH "Adolescence+") OR (MH "Young Adult") OR (MH "Child")) AND (MH “Indigenous Peoples+”)) | 5,431 |
| **#3** | S1 OR S2 | 9,555 |
| **#4** | (MH "Canada+") OR (MH "Arctic Regions") | 106,724 |
| **#5** | TX (Arctic OR circumpolar OR “Newfoundland and Labrador” OR “Nova Scotia*” OR Canadian* OR Canada OR “New Brunswick” OR Quebec* OR Ontario OR Manitoba* OR Saskatchewan OR Alberta* OR “British Columbia*” OR “B.C.” OR “Northwest Territories” OR “N.W.T.” OR Nunavut OR “Yukon Territory”) | 509,284 |
| **#6** | S4 OR S5 | 509,311 |
| **#7** | (MH "Suicide+") OR (MH "Mental Health") OR (MH "Mental Disorders+") OR (MH "Adolescent Psychology") OR (MH "Child Psychology") OR (MH "Psychology, Social+") OR (MH "Psychology") OR (MH "Hardiness") OR (MH "Community Mental Health Services") OR (MH "Cultural Competence”) OR (MH "Health Services Accessibility+") OR (MH "Health Services+") OR (MH "Substance Use Disorders+") OR (MH "Child Psychiatry") OR (MH "Behavioral Symptoms") | 2,437,255 |
| **#8** | TI (“mental* ill*” OR suicid* OR resilen* OR “protective factors” OR “mental health” OR coping OR cope OR “mental wellness” OR “mental wellbeing” OR “psycho-social” OR psychosocial OR “cultural safety” OR “cultural competence” OR “social support” OR trauma* OR “cultur* connectedness” OR “mental toughness” OR adaptability OR “community involvement” OR “community support*” OR “psychosis” OR “mental disorder*” OR “substance-related disorder*” OR “substance abuse” OR “substance use*” OR “drug use*” OR “drug abuse” OR “drug dependence” OR alcoholism OR “alcohol abuse” OR “disruptive behavior*” OR “disruptive behaviour*” OR "Addiction" OR "Alcohol Use" OR "Cannabis Use*" OR “cannabis abuse” OR "Drug Dependenc*" OR "Inhalant Abuse" OR “inhalant use*” OR “opioid abuse” OR "Opioid Use*" OR "Tobacco Use*” OR “prescription abuse” OR “chronic stress” OR “acute stress” OR “adjustment disorder*” OR “stress disorder*” OR depression OR “depressive symptoms”)  OR AB (“mental* ill*” OR suicid* OR resilen* OR “protective factors” OR “mental health” OR coping OR cope OR “mental wellness” OR “mental wellbeing” OR “psycho-social” OR psychosocial OR “cultural safety” OR “cultural competence” OR “social support” OR trauma* OR “cultur* connectedness” OR “mental toughness” OR adaptability OR “community involvement” OR “community support*” OR “psychosis” OR “mental disorder*” OR “substance-related disorder*” OR “substance abuse” OR “substance use*” OR “drug use*” OR “drug abuse” OR “drug dependence” OR alcoholism OR “alcohol abuse” OR “disruptive behavior*” OR “disruptive behaviour*” OR "Addiction" OR "Alcohol Use" OR "Cannabis Use*" OR “cannabis abuse” OR "Drug Dependenc*" OR "Inhalant Abuse" OR “inhalant use*” OR “opioid abuse” OR "Opioid Use*" OR "Tobacco Use*” OR “prescription abuse” OR “chronic stress” OR “acute stress” OR “adjustment disorder*” OR “stress disorder*” OR depression OR “depressive symptoms”) | 592,034 |
| **#9** | S6 OR S7 | 2,752,063 |
| **#10** | S3 AND S6 AND S9 | 2,003 |

**PsycInfo**

Searches conducted on August 26, 2021

| **Search** | **Query** | **Records Retrieved** |
| --- | --- | --- |
| #1 | (DE "Adolescent Development" OR DE "Adolescent Behavior" OR DE “Emerging Adulthood” OR DE "Early Adolescence") AND (DE "Indigenous Populations" OR DE "Inuit”) | 123 |
| #2 | TI (child* OR youth* OR adolescent* OR “school age” OR “middle school” OR “junior high” OR “high school” OR teen* OR “young adult” OR “emerging adult”) AND (inuit OR metis OR aboriginal* OR indigenous OR “first people” OR “first nation*” OR native)  OR AB (child* OR youth* OR adolescent* OR “school age” OR “middle school” OR “junior high” OR “high school” OR teen* OR “young adult” OR “emerging adult”) AND (inuit OR metis OR aboriginal* OR indigenous OR “first people” OR “first nation*” OR native) | 11,751 |
| #3 | S1 OR S2 | 11,756 |
| #4 | TX (“Arctic Regions” OR arctic OR circumpolar OR “Prince Edward Island” OR “P.E.I.” OR “Newfoundland and Labrador” OR “Nova Scotia*” OR Canadian* OR Canada OR “New Brunswick” OR Ontario OR Manitoba* OR Saskatchewan OR Alberta* OR “British Columbia*” OR “B.C.” OR “Northwest Territories” OR “N.W.T.” OR “Nunavut” OR “Yukon Territory”) | 347,047 |
| #5 | ((((((((((DE "Suicidal Behavior" OR DE "Youth Suicide" OR DE "Mental Disorders" OR DE "Self-Injurious Behavior" OR DE "Suicide Prevention") OR (DE "Mental Health")) OR (DE "Psychology")) OR (DE "Adolescent Psychology")) OR (DE "Resilience (Psychological)")) OR (DE "Emotional Adjustment")) OR (DE "Adaptability (Personality)")) OR (DE "Community Mental Health Services")) OR (DE "Cultural Competence")) OR (DE "Health Care Access")) OR (DE "Mental Health Services" OR DE "Substance Use Disorder" OR DE "Addiction" OR DE "Alcohol Use Disorder" OR DE "Cannabis Use Disorder" OR DE "Drug Abuse" OR DE "Drug Dependency" OR DE "Inhalant Abuse" OR DE "Opioid Use Disorder" OR DE "Tobacco Use Disorder" OR DE "Child Psychiatry" OR DE "Acute Psychosis" OR DE "Acute Schizophrenia") | 399,746 |
| #6 | TI (“mental* ill*” OR suicid* OR resilen* OR “protective factors” OR “mental health” OR coping OR cope OR “mental wellness” OR “mental wellbeing” OR “psycho-social” or psychosocial OR “cultural safety” OR “cultural competence” OR “social support” OR trauma* OR “cultur* connectedness” OR “mental toughness” OR adaptability OR “psychosis” OR “mental disorder*” OR “substance-related disorder*” OR “substance abuse” OR “substance use*” OR “drug use*” OR “drug abuse” OR “drug dependence” OR alcoholism OR “alcohol abuse” OR “disruptive behavior*” OR “disruptive behaviour*” OR "Addiction" OR "Alcohol Use" OR "Cannabis Use*" OR “cannabis abuse” OR "Drug Dependenc*" OR "Inhalant Abuse" OR “inhalant use*” OR “opioid abuse” OR "Opioid Use*" OR "Tobacco Use*” OR “prescription abuse” OR “chronic stress” OR “acute stress” OR “adjustment disorder*” OR “stress disorder*” OR depression OR “depressive symptoms”)  OR AB (“mental* ill*” OR suicid* OR resilen* OR “protective factors” OR “mental health” OR coping OR cope OR “mental wellness” OR “mental wellbeing” OR “psycho-social” or psychosocial OR “cultural safety” OR “cultural competence” OR “social support” OR trauma* OR “cultur* connectedness” OR “mental toughness” OR adaptability OR “community involvement” OR “community support*” OR “psychosis” OR “mental disorder*” OR “substance-related disorder*” OR “substance abuse” OR “substance use*” OR “drug use*” OR “drug abuse” OR “drug dependence” OR alcoholism OR “alcohol abuse” OR “disruptive behavior*” OR “disruptive behaviour*” OR "Addiction" OR "Alcohol Use" OR "Cannabis Use*" OR “cannabis abuse” OR "Drug Dependenc*" OR "Inhalant Abuse" OR “inhalant use*” OR “opioid abuse” OR "Opioid Use*" OR "Tobacco Use*” OR “prescription abuse” OR “chronic stress” OR “acute stress” OR “adjustment disorder*” OR “stress disorder*” OR depression OR “depressive symptoms”) | 943,972 |
| #7 | S5 OR S6 | 1,076,814 |
| #8 | S3 AND S4 AND S7 | 619 |

**ERIC**

Searches conducted on August 26, 2021

| Search | Query | Results Retrieved |
| --- | --- | --- |
| #1 | (MAINSUBJECT.EXACT("Late Adolescents") OR MAINSUBJECT.EXACT("Early Adolescents") OR MAINSUBJECT.EXACT("Preadolescents") OR MAINSUBJECT.EXACT.EXPLODE("Youth") OR MAINSUBJECT.EXACT.EXPLODE("Secondary School Students") OR MAINSUBJECT.EXACT("Adolescents") OR MAINSUBJECT.EXACT("Young Adults")) AND (MAINSUBJECT.EXACT("Canada Natives") OR MAINSUBJECT.EXACT("Eskimos") OR MAINSUBJECT.EXACT("Indigenous Populations")) | 615 |
| #2 | ti ( (child* OR youth* OR adolescent* OR “school age” OR “middle school” OR “junior high” OR “high school” OR teen* OR “young adult” OR “emerging adult”) AND (“first nations” OR “inuit” OR metis OR aboriginal* OR indigenous OR “first people” OR native) )  OR ab ( (child* OR youth* OR adolescent* OR “school age” OR “middle school” OR “junior high” OR “high school” OR teen* OR “young adult” OR “emerging adult”) AND (“first nations” OR “inuit” OR metis OR aboriginal* OR indigenous OR “first people” OR native)) | 8,635 |
| #3 | 1 OR 2 | 8,913 |
| #4 | Canada OR Canadian* OR “Arctic Regions” OR arctic OR circumpolar OR “Prince Edward Island” OR “P.E.I.” OR “Newfoundland and Labrador” OR “Nova Scotia*” OR “New Brunswick” OR Quebec* OR Ontario OR Manitoba* OR Saskatchewan OR Alberta* OR “British Columbia*” OR “B.C.” OR “Northwest Territories” OR “N.W.T.” OR Nunavut OR “Yukon Territory” | 65,760 |
| #5 | MAINSUBJECT.EXACT("Coping") OR MAINSUBJECT.EXACT.EXPLODE("Mental Disorders") OR MAINSUBJECT.EXACT("Resilience (Psychology)") OR MAINSUBJECT.EXACT("Access to Health Care") OR MAINSUBJECT.EXACT("Mental Health") OR MAINSUBJECT.EXACT("Suicide") OR MAINSUBJECT.EXACT("Child Psychology") OR MAINSUBJECT.EXACT("Community Health Services") OR MAINSUBJECT.EXACT.EXPLODE("Substance Abuse") OR MAINSUBJECT.EXACT.EXPLODE("Child Psychology") OR MAINSUBJECT.EXACT.EXPLODE("Psychosis") | 78,985 |
| #6 | ti (“mental* ill*” OR suicid* OR coping OR cope OR resilen* OR “protective factors” OR “mental health” OR “mental wellness” OR “mental wellbeing” OR “psycho-social” OR psychosocial OR “cultural safety” OR “cultural competence” OR “social support” OR “community mental health services” OR “culturally competent care” OR “health services” OR “health services accessibility” OR trauma* OR “cultur* connectedness” OR “mental toughness” OR adaptability OR “community involvement” OR “community support*” OR “psychosis” OR “mental disorder*” OR “substance-related disorder*” OR “substance abuse” OR “substance use*” OR “drug use*” OR “drug abuse” OR “drug dependence” OR alcoholism OR “alcohol abuse” OR “disruptive behavior*” OR “disruptive behaviour*” OR "Addiction" OR "Alcohol Use" OR "Cannabis Use*" OR “cannabis abuse” OR "Drug Dependenc*" OR "Inhalant Abuse" OR “inhalant use*” OR “opioid abuse” OR "Opioid Use*" OR "Tobacco Use*” OR “prescription abuse” OR “chronic stress” OR “acute stress” OR “adjustment disorder*” OR “stress disorder*” OR depression OR “depressive symptoms”)  OR ab (“mental* ill*” OR suicid* OR coping OR cope OR resilen* OR “protective factors” OR “mental health” OR “mental wellness” OR “mental wellbeing” OR “psycho-social” OR psychosocial OR “cultural safety” OR “cultural competence” OR “social support” OR “community mental health services” OR “culturally competent care” OR “health services” OR “health services accessibility” OR trauma* OR “cultur* connectedness” OR “mental toughness” OR adaptability OR “community involvement” OR “community support*” OR “psychosis” OR “mental disorder*” OR “substance-related disorder*” OR “substance abuse” OR “substance use*” OR “drug use*” OR “drug abuse” OR “drug dependence” OR alcoholism OR “alcohol abuse” OR “disruptive behavior*” OR “disruptive behaviour*” OR "Addiction" OR "Alcohol Use" OR "Cannabis Use*" OR “cannabis abuse” OR "Drug Dependenc*" OR "Inhalant Abuse" OR “inhalant use*” OR “opioid abuse” OR "Opioid Use*" OR "Tobacco Use*” OR “prescription abuse” OR “chronic stress” OR “acute stress” OR “adjustment disorder*” OR “stress disorder*” OR depression OR “depressive symptoms”) | 85,642 |
| #7 | 5 OR 6 | 126,622 |
| #8 | 3 AND 4 AND 7 | 153 |

**Scopus**

Searches conducted on August 26, 2021

| Search | Query | Results Retrieved |
| --- | --- | --- |
| #1 | TITLE-ABS-KEY ( (Adolescent* OR “young adult” OR child OR children OR youth* OR “school age” OR “middle school” OR “junior high” OR “high school” OR teen* OR “young adult” OR “emerging adult”) AND (“Indigenous Canadian*” OR inuit OR metis OR “first nations” OR aboriginal* OR indigenous OR “first people” OR native) ) | 42,666 |
| #2 | TITLE-ABS-KEY ( Canada OR Canadian* OR “Arctic regions” OR Arctic OR {Prince Edward Island} OR “Newfoundland and Labrador” OR "Nova Scotia*" OR {P.E.I.} OR {New Brunswick} OR Quebec* OR Ontario OR Manitoba* OR Saskatchewan OR Alberta* OR “British Columbia*” OR {B.C.} OR “Northwest Territories” OR {N.W.T.} OR Nunavut OR “Yukon Territory” ) | 725,032 |
| #3 | TITLE-ABS-KEY (“mental* ill*” OR Suicid* OR {mental health} OR “mental disorder*” OR {adolescent psychology} OR {psychological resilience} OR resilen* OR “protective factors” OR coping OR cope OR {mental wellness} OR “mental wellbeing” OR {psycho-social} OR psychosocial OR “cultural safety” OR “cultural competence” OR “social support*” OR “community mental health services” OR “culturally competent care” OR “health services” OR “health services accessibility” OR trauma* OR “cultur* connectedness” OR {mental toughness} OR adaptability OR “community involvement” OR “community support*” OR {Behavioral Symptoms} OR {Adolescent Psychiatry} OR {Substance-Related Disorders} OR “psychosis” OR “mental disorder*” OR “substance-related disorder*” OR “substance abuse” OR “substance use*” OR “drug use*” OR “drug abuse” OR “drug dependence” OR alcoholism OR “alcohol abuse” OR “disruptive behavior*” OR “disruptive behaviour*” OR “Addiction” OR “Alcohol Use” OR “Cannabis Use*” OR “cannabis abuse” OR “Drug Dependenc*” OR “Inhalant Abuse” OR “inhalant use*” OR “opioid abuse” OR “Opioid Use*” OR “Tobacco Use*” OR “prescription abuse” OR “chronic stress” OR “acute stress” OR “adjustment disorder*” OR “stress disorder*” OR depression OR “depressive symptoms” ) | 3,570,197 |
| #4 | #1 AND #2 AND #3 | 1,393 |
