## Supplementary File 2 for "Resilience and Protective Factors for Mental Health among Indigenous Youth in Canada: A Scoping Review"

**Data Extraction Instrument**

| **General characteristics** | |
| --- | --- |
| Study title |  |
| Authors |  |
| Year of publication |  |
| Literature type |  |
| Country of publication |  |
| Research aim/purpose/objective |  |
| Methodology/methods |  |
| Geographical setting (province(s), region)  (Where the study is taking place) |  |
| Type of service  (healthcare, education, community) |  |
| Description of community involvement |  |
| Involved community, knowledge users, or stakeholders | With whom did the trainee partner with?  e.g., patients, health care providers, policymakers, government, etc. |
| Use of a theory, model, or framework (TMF) | NR if no TMF |
| **Resilience/Protective Factors for mental health** | |
| Definition of resiliency/protective factors/coping skills used |  |
| Identified resiliency/protective |  |
| Description of resilience or protective factors |  |
| Type of mental health studied |  |
| **Indigenous youth characteristics** | |
| Study population (participants) inclusion criteria (under methods section usually) |  |
| Reported descriptions (e.g., age, gender, demographics)  Read the results section |  |
| Indigenous community names |  |
| **Outcome** | |
| Outcome(s) of interest |  |
| Outcome measurement tool(s) |  |
| Reported outcomes |  |
| Study main findings and conclusion |  |
| Future implications |  |
| Other notes |  |
