## Supplementary File 3 for "Resilience and Protective Factors for Mental Health among Indigenous Youth in Canada: A Scoping Review"

| **Author, year** | **Literature Type** | **Research Aim/Purpose/Objective** | **Design** | **Geographical Setting** | **Type of mental health concern** | **Study participants** | **Indigenous communities** | **Outcome(s) of interest** |
| --- | --- | --- | --- | --- | --- | --- | --- | --- |
| Adams 2015 | Peer-reviewed | Examine how an Equine Assisted Learning (EAL) program contribute to the wellbeing of First Nations female youth who misuse volatile substances? | Case study | Saskatchewan, Calgary, Alberta and Regina | Substance use | First Nations, female youth, 12–18 years of age who misuse volatile substances | NR | Impact of the Equine Assisted Learning (EAL) program on First nations youth's wellbeing: Biological or physical wellbeing, Psychological (mental/emotional) wellbeing, Social Wellbeing, Spiritual wellbeing |
| Ames 2015 | Peer-reviewed | Describe the developmental trajectory of depressive symptoms with an additional goal of identifying gender differences in depressive symptoms.  Examine the relationship between alcohol use behaviours, self-esteem, optimism, and the trajectories of depressive symptoms. Determine whether self-esteem and optimism mediated the relationship between alcohol use and subsequent depressive symptoms. | Observational | Canada (national) | Depression, substance | Off-reserve Aboriginal youth were selected from cycles 4–7 of the National Longitudinal Survey of Children and Youth. Mean Age:12, Gender: 48.3% male and 51.7% female, Household income; Under $20,000: 14.4%, Between $20,000 and $40,000: 28.9% and Over $40,000: 56.6 Ethnic origin; North American Indian 67.6%, Metis: 23.9% and Other (e.g., Inuit): 8.5% | Metis, Inuit | Depressive Symptoms, Alcohol use, Self-esteem, Optimism |
| Ansloos 2021 | Peer-Reviewed | Present findings from research with a group of 15 Indigenous youth leaders working in community health, suicide prevention and mental health organizations across Canada to understand how they are conceptualizing mental health and its intersection with spirituality. | Qualitative | Ontario, Canada | Suicide, stress | Disclosure of participant’s gender, sexuality, age, and other demographic information was not solicited, beyond what participants volunteered on their own accord without prompting. | First Nations, Inuit, Metis | Indigenous young peoples’ conceptualizations of mental health and wellness |
| Arato-Bollivar 2005 | Grey Lit | Develop categories that would identify themes in the events reported by Aboriginal individuals, by exploring the research question: "What are the critical incidents contributing to survival in suicidal aboriginal youth?" | Qualitative | British Columbia, Canada | Suicide | 20 participants ranged in age from 18 to 61. Mean age = 37.5. The majority of the participants were in their mid-30s to 40s. Geographically, all individuals were born and grew up in British Columbia with 18 currently living in a small Northern community in BC. The majority of the participants identified both parents with Aboriginal origin. Female (n=14) and male (n=6). | First Nations | Survival factors present in suicidal aboriginal youth |
| Feathers of Hope 2014 | Grey lit | Develop the action plan as a five-year road map that we believe will move forward the vision of young people and provide a real opportunity for youth to be part of shaping the work tied to the healing processes in our individual communities. | Qualitative | Thunder Bay, Ontario & Kashechewan/Ft. Albany | Substance use, suicide, isolation, trauma | First Nations youth from 92 communities in Ontario | First Nations | Understand the issues facing First Nations Indigenous youth in remote communities in Ontario and how they feel these issues can be addressed using the action plan. |
| Baydala 2014 | Peer-reviewed | Review and cultural adaptation of the elementary and junior high Life Skills TrainingLST programs, delivery of the adapted programs, measurement of changes in student knowledge of the negative effects of drug and alcohol use, attitudes toward drug and alcohol use, drug and alcohol refusal skills, and changes in self-esteem/self-concept, and documentation of the community’s experience of and responses to the program adaptation and delivery. | Participatory Action Research | Alberta | Substance | Children in grades 3, 4, 5, 6, 7, 8 at Alexis Nakota Sioux Nation School | First Nations | Effectiveness of the program (experience), and pre- post participants’ Knowledge and behaviour (i.e., health knowledge, attitudes, drug refusal skills, and self-esteem). |
| Bohr 2016 | Peer-reviewed abstract | Overview of a pilot trial of computer-based e-therapy offered to youth in remote northern communities who are at risk for depression and suicidality. | Experimental | Nunavat | Depression | 5 youth in 25 communities in Northern Canada | Inuit | Depressive symptoms and resilience |
| Chandler 1998 | Peer-reviewed | Examine self-continuity and its role as a protective factor against suicide. | Observational | British Columbia | Suicide | N/A | NR | Suicide rates, sociodemographic data, markers of attempted cultural rehabilitation |
| Crooks 2010 | Peer-reviewed | Develop strength-based violence prevention programs for First Nations youth. The primary objective is to promote healthier relationships and develop youth leadership skills to increase engagement and school connectedness. The initiatives described include a peer mentoring program, a First Nations Cultural Leadership Course, and Grade 8 Transitions Conferences. | Participatory Action Research | London, Ontario, Canada | Substance | First Nations students in grades 8 and 9 in the Thames Valley District School Board | First Nations | Youth engagement in school and community |
| Crooks and Dunlop 2017 | Peer-reviewed | Evaluate impacts of two years of program participation on positive well-being and cultural identity, across the transition from elementary to secondary school. | Case study | Ontario | General mental health and well-being | 82 individuals, including elementary and secondary students as well as five educators and two administrators from 15 schools. | Inuit, First nations | Cultural identity and mental health outcomes |
| Crooks 2017 | Peer-reviewed | Longitudinal evaluation of the effects of 1 or 2 years of program participation on positive well-being, as assessed by mental health and cultural identity, across the transition from elementary to secondary school. | Mixed methods | Southwestern Ontario | General mental health and well-being | 105 students, Grade 7 and 8. Age: 12.59 (11-14); sex: Male 50.5(53), Female 49.5(52); Grade 7: 39 (41); Grade 8: 61 (64) | First Nations, Metis, Inuit | Mental health outcomes, cultural identify, school climate life satisfaction |
| Dell 2011 | Peer-reviewed | Understand the experiences of First Nations and Inuit youth participating in an EAL program as part of their healing from solvent abuse while in a residential treatment centre. | Qualitative | Muncey, ON | Substance | 15 youth participants: male (n=7) and female (n=8). The male participants ranged from 12 to 17 years of age, with an average age of 14. Five of the male youth were of First Nations descent from five different cultural territories in Canada, and two were Inuit from one territory. All youth were at the NNHC for solvents as their primary substance of abuse, although 86% also problematically used alcohol and other drugs. Only two male youth had prior experience with horses, and the two Inuit youth had never seen a living horse before. The average age of the female participants was 15, and their age ranged from 13 to 17. Of the eight girls, six were First Nations, and similar to the males, represented six diverse cultural territories in Canada. Also similar to the boys, all girls were attending NNHC for solvent abuse; however, 88% also abused other substances. Three female youth had prior exposure to horses. | First Nations + Inuit | Impact of Equine Assisted Learning |
| Dell and Hopkins,2011 | Peer-reviewed | Illustrate the role of Indigenous culture and its intersection with Western approaches to recovery in the operation of Youth Solvent Addiction Program (YSAP)’s residential treatment centers | Multi-methods Mixed | Saskatchewan | Substance | A sample of youth who entered the nine centers between 2006 and 2008 (N=267,37% male and 63% female). First Nations and Inuit youth ranging in age from 12 to 26. | Anishinaabe | Completion of treatment at a YSAP center |
| Dunlop 2016 | Thesis | Examine the bullying experiences of FNMI youth, the effects of these experiences on mental health and well-being, and the potential moderating effect of three protective factors (cultural, school, and peer connectedness), using longitudinal data collected from a cohort of FNMI adolescents in a large school district in southwestern Ontario. | Observational | Southwestern Ontario | General mental health and well-being | Youth aged 11-14 years, in grades 7 to 10, self-identified as First Nations, Inuit, or Metis | First Nations, Metis, and Inuit | Bullying victimization and perpetration, positive mental health and well-being, adolescent connectedness, cultural connectedness |
| Eggertson 2013 | News article - grey lit | Highlight the importance of the Applied Suicide Intervention Skills Training program for people in First Nations community | N/A | Nunavut | Suicide | NR | First Nations | Completion of Applied Suicide Intervention Skills Training (ASIST) training in mental health crisis |
| Fanian 2015 | Peer-reviewed | Evaluate a creative arts workshop for Tłı˛cho˛ youth where youth by exploring critical community issues and find solutions together using the arts. Develop a community-led, youth-driven model to strengthen resiliency through youth engagement in the arts in circumpolar regions. | Mixed methods | Northwest Territories | General mental health and well-being | All youth were Tłı ̨cho ̨and from the community of Behchoko` ̨, NT. The youths’ ages ranged from 13 to 22; there were 4 females and 5 males overall. Five Indigenous artist facilitators delivered the creative arts programming. | Tłı˛cho˛ | Youth perspective, art products, facilitator perspectives |
| Filbert 2010 | Peer-reviewed | Test the hypothesis that among First Nations young people in care, develop-mental and cultural assets would be associated with more resilient mental health and educational outcomes. | Observational | Ontario | General mental health and well-being | Drawn from a larger OnLAC sample of young people who were in care and aged 10–17 in 2005–2006, the First Nations sample consisted of 97 children and adolescents (49males and 48 females [M=12.96 years of age, SD=2.00]). The vast majority of the participants were living in foster care (97%), with a few in kinship care (3%). | NR | Mental health (prosocial behaviour and self-esteem), resilience (educational performance and difficulty scale) |
| Filbert 2014 | Grey Lit | Examine resilience-promoting factors on the child, family, and community levels among Aboriginal young people in care. | Mixed methods | Ottawa, Ontario | Suicide depression | First study: 510 First Nations (237 females, 273 males aged 10-16 years), 39 Métis (15 females, 24 males aged 10-16 years), and 10 Inuit young people (2 females, 8 males aged 10-16 years).  The second study: 21 First Nations children and adolescents residing in out-of-home care in northern Ontario. | First Nations, Metis, and Inuit | Study 1 1: Health, education, identity, family and social relationships, social presentation, emotional and behavioural development, and self-care skills 2: Identification with culture 3: Negative mental health 4: Goal accomplishment 5: Developmental Assets 6: Positive mental health  7: Educational performance demographics: Age, ethnicity, placement type  Study 2:  General resilience questions, in-depth interview questions (i.e., support, empowerment, boundaries and expectations, constructive use of time, commitment to learning, positive values, social competencies, positive identity) |
| First Nations Information Governance Centre 2021 | Report | Identify potential protective factors against cigarette smoking among First Nations youth living on reserves and in northern communities. | Mixed-methods | Reserves and northern communities across Canada | Substance use | 3,842 youth, aged 12 to 17 | First Nations | Youth smoking behaviours |
| First Nations Information Governance Centre 2014 | Grey lit | Explore the concept of resilience in the context of youth suicide and negative well-being. | Observational | Alberta, Manitoba, Ontario and Quebec | Depression, suicide, substance | 4,837 First Nations youth aged 12-17 years | First Nations | Suicidal ideation and/or attempts, protective factors exist in communities. |
| Flanagan 2011 | Peer-reviewed | The author's primary hypothesis was that a strong cultural identity would predict less perceived physical and relational aggression. | Observational | Kawawachikamach, Quebec | Depression, anxiety | Students enrolled in grades 6 and 11, have one parent of Indigenous descent. The data of the 62 (36 male) students who identified one or both of their parents as Aboriginal (Montagnais or Naskapi) and who completed most of the questionnaires were included in the analyses. Of which, 54 of these students identified themselves as Aboriginal, and 8 others identified themselves as white and Aboriginal. Mean age= 13.7 years, with a range of 11–19 years. All the adolescents had one or both Naskapi parents. | Innu | Self-reports: cultural identity and internalizing behaviour (depression and anxiety), Peer ratings: physical aggression, relational aggression, and prosocial behaviour and Teacher ratings |
| Fraser 2015 | Peer-reviewed | Explore the prevalence of suicide ideations and attempts among 15–24-year-olds living in Nunavik, Quebec, and to explore the risk and protective factors of suicide attempts as a function of gender. | Observational | Nunavik, Quebec | Depression, suicide | Permanent residents of Nunavik excluding residents of collective dwellings and households in which there were no Inuit aged 18 years and over. A total of 305 Inuit between the ages of 15 and 24 completed the survey. | Inuit | Psychological distress, alcohol misuse, substance use, interpersonal violence, potential protective factors with gender differences, and their impacts on suicidal attempts/thoughts |
| Gfellner 2016 | Peer-reviewed | Investigate associations between ego strengths and racial/ethnic identity and personal adjustment/well-being among 178 North American Indian/First Nations adolescents who resided and attended school on reserves. | Observational | Southern Midwest of Canada | Stress | 178 adolescents from First Nations Communities, in grades 7 and 8: Females (n=39), Males (n=42) Grades 9 to 12: Females (n=50), Males (n=47) | First Nations | Well-being, ego strengths, self-esteem, adjustment |
| Goldstein 2021 | Peer-reviewed | Examine the moderating role of valuing cultural activities on the relationship between positive alcohol expectancies and alcohol use and heavy drinking in a sample of Indigenous youth. | Observational | A First Nations reserve in Eastern Canada | Substance use | 106 First Nation adolescents from eastern Canadian reserve communities, ranged in age from 11 to 18 and were 50% female. | First Nations | Alcohol expectancies, alcohol use, heavy drinking, cultural activities |
| Hackett 2016 | Peer-reviewed | Going Off, Growing Strong is a program for Inuit youth facing widespread social, cultural, and economic change. The overarching goals of the program are to enhance resilience and wellness; build social connections for the youth; and transmit traditional knowledge, skills, and values to participating youth | Participatory Action Research | Ontario | Suicide | School-age Inuit males in Nain, Nunatsiavut considered at risk for suicidal behaviors | Inuit | Suicide |
| Hallett 2007 | Grey lit | Test the hypothesis that community-level efforts to contribute to the preservation of indigenous languages constitute an additional marker of cultural continuity that will again prove to be strongly and independently associated with youth suicide rates. | Observational | British Columbia | Suicide | NR | NR | Suicide rates; total number of cultural factors present |
| Hammond 2000 | Grey lit | Examine the perceived patterns of attachment of three naturally occurring groups of Native adolescents-56 solvent users, 80 poly-substance users, and 88 nonsubstance users - their attachment relationships to their parents and peers as well as to explore their perception of well-being and social adaptation based on early experiences with attachment figures. | Observational | Calgary | Substance use | 224 Native adolescent volunteers in the age range of 12-18 years. mean age 14.69 years; females (n=98), males (n=126). 41% biologically intact family 34% single parent  21% blended 2.7% not residing with either parent | NR | Attachment characteristics, perception of well-being and social adaptation characteristics |
| Hatala 2017 | Peer-Reviewed | Demonstrate how concepts of time and the future inform processes of resilience among Indigenous adolescents within an urban Canadian context | Qualitative | Saskatoon | Substance, suicide | 28 Indigenous youth (ages 15–25) Plains Cree (21) and Metis (7) youth aged 15-25, 12 male and 16 female. 8 attending high school, 5 with post-secondary education, 13 working/not in school, 9 parents | Cree, Metis | Resilience strategies, motivations for health and wellness-seeking behaviors, belonging, self-mastery, cultural identity |
| Hirsch 2016 | Peer-reviewed | Overview of background to intergenerational healing initiatives in Nunatsiavut; our interpretive approach; community context; objectives of the Going Off, Growing Strong program; roles of program staff; youth activities; and relationships between program objectives, activities, and community resilience. | N/A | Nain, Nunatsiavut -(Labrador, Canada) | Stress, anxiety, stress | Inuit male youth from Nain | Inuit | Providing template for co-creating a conceptual framework and resulting outcome measures, so that they can better understand the impact of such a program on chosen outcomes |
| Hutt-MacLeod 2021 | Peer-reviewed | Describe the implementation of the ACCESS Open Minds (ACCESSOM) objectives for youth mental health service transformation within a pre-existing Fish Net Model of transformative youth mental healthcare service in the community of Eskasoni. | Participatory Action Research | Cape Breton Island, Nova Scotia | Substance, suicide | NR | First Nations | Early case identification, rapid access to initial assessment, availability of appropriate services, engagement of youth and families, and elimination of transitions in service based on age. |
| Isaak 2020 | Peer-reviewed | Explore the fit between on-reserve First Nations community members’ conceptualizations of help-seeking for mental health concerns and the Andersen Behavioral Model of Health Services Use. | Participatory Action Research | Northwestern/central Regions of Canada | General mental health and well-being | There were 79 (69%) females and 36(31%) males. Participants included 24 youth and 97 adults, including three elders, age range of 13 to 80 years, with 82%) self-identifying as indigenous Cree descent. The median age is between 17.2 and 22.0 years | First Nations, Cree, Metis, Ojibwe, Oji-Cree, Plains Ojibwe | Degree of fit between FN community members’ conceptualizations of help-seeking for mental health concerns and four ABM domains. Family, community and informal and formal support contexts and systems in conjunction with pathways to care for mental health problems |
| Ivanich 2020 | Peer-reviewed | Provide two case studies of substance use prevention programs, carried out by independent research teams with long partnerships with distinct Indigenous populations. Case 1- Canada | Participatory Action Research | Case study 1: upper Midwest and Canada | Substance use | 5th-8th grade adolescents and their families, 3rd and 4th graders | Anishinaabe (Ojibwe) | Impacts on substance use prevention, delay of onset, and promoting the wellbeing of Indigenous youth and families. |
| Janelle 2009 | Peer-reviewed | Aimed at increasing self-esteem by exposing the teenagers to demanding situations in which they must mentally and physically out-perform themselves. Second, through the teachings of traditional practices and the use of the vernacular language, the project proposes to reinforce culture and encourage the use of pro-social behaviour among participants through cooperation and mutual help. Finally, the project aims to mobilise the community and encourage families to get actively involved. | Mixed methods | Atikamek communityof Manawan Quebec | Suicide | Six 14 to 17-year-old males from the Manawan community. | Atikamekw First Nation | Situational self-esteem, dispositional self-esteem, implementation, pro-social behaviour, cultural pride, cultural promotion community mobilization |
| Katapally 2020 | Peer-reviewed | Describes evidence-based strategies to overcome barriers to implementation through the integration of citizen science and community-based participatory research action. | N/A - opinion piece | Saskatchewan, Canada | General mental health and well-being + substance | 34 youth citizen scientists (n=16 from School 1, n=18 from School 2) | First Nations, Inuit, and Metis | Benefits of program, barriers to implementation, evidence-based strategies to address barriers |
| Kirmayer 1998 | Peer-reviewed | Identify potential risk and protective factors associated with attempted suicide among Inuit youth, a population known to have a high rate of both attempted and completed suicide in recent years | Observational | Sante Quebec | Suicide, substance, mental illness | Inuit youth (aged 15 to 24years) from a random community survey conducted by Sante Quebec in 199 | Inuit | Psychiatric illness, suicide, alcohol abuse |
| Kral 2014 | Peer-reviewed | Explore the question of how Inuit youth cope with the difficulties they experience in their lives. We examined personal, relational, and community levels of coping and resilience | Qualitative | Igloolik, Nunavut, Canada | General mental health and well-being | 23 youth participants, aged 12–19, from the elementary and high schools of the community. | Inuit | life experiences, stressors, and coping or resilience strategies |
| Liebenberg 2022 | Peer-reviewed | Report on findings from a Participatory Action Research (PAR) study conducted in a First Nations community in Unama’ki (Cape Breton), Atlantic Canada. | Participatory Action Research | Unama’ki (Cape Breton), Nova Scotia, Atlantic Canada | General mental health and well-being | Eight first nations youth participants (14–18 years old) from the Eskasoni First Nations community | Mi'kmaq | Indigenous youth engagement and resiliency by drawing on relational (space-based) and contextual (place-based) resources "Spaces & Places" |
| Linds 2019 | Peer-reviewed | Describe the process and outcomes of a project that taught film and film production in a First Nations high school located in a predominantly Neehithuw community in northern Saskatchewan. Within the context of a holistic view of health and wellness among Indigenous youth, the article highlights the content of the films, the rewards of the filmmaking experience, the challenges faced by the students during the process, and the teacher’s reflections on the process. | Participatory Action Research | Neehithuw, Northern Sasketwchan | General mental health and well-being | Grade 10 Communications Media course at a First Nations high school in a Neehithuw (Woodland Cree) community in northern Saskatchewan. | Cree | Review of three fictional films, examined the process of making the films using primarily the documentary and the answers to the questionnaire. |
| Litwin 2023 | Peer-reviewed | Build knowledge with Inuit youth about their own community’s cultural fit with Smart, Positive, Active, Realistic, X-Factor (SPARX) and to understand the possible benefits and drawbacks of SPARX, as determined by youth and community facilitators in Nunavut. | Qualitative | Nunavut | General mental health and well-being | 11 youth participants were between the ages of 13–18 and had been identified by the community facilitators as exhibiting low mood, negative affect, depressive presentations, and/or significant levels of stress. | Inuit | Experiences using SPARX |
| Lys 2018 | Peer-reviewed | Explore the self-identified strategies that female youth in the NWT use to cope with mental health issues. | Participatory Action Research | the Northwest Territories, Canada, | Post-traumatic stress disorder, depression, bipolar, anxiety, suicide, substance, trauma | Females 13-18 years old, Aboriginal or indigenous (First Nations, Metis, or Inuit) | First Nations, Metis, Inuit | The personal narratives of the body map |
| MairTiessen 2008 | Grey Lit | Investigated the impact of collective control and identity on the well-being of Aboriginal youth | Mixed methods | Manitoba | Substance use | 82 youth from two Cree communities in northern Manitoba participated; 52 female and 30 males. | Cree | Individual-level control, Group-level control, cultural affinity, self-esteem, affect, general happiness, substance abuse |
| Miller 2011 | Peer-reviewed | Enrich culturally the FRIENDS for Life program, an evidence-based curriculum for anxiety, with culturally sensitive Aboriginal content, and prevent and reduce anxiety symptoms in children by offering the enriched FRIENDS for Life program in public school classroom contexts. | Quasi experimental | Western Canada | Anxiety | 192 Aboriginal students. Boys and girls were equally represented in the sample (50.1% girls), with a mean age of 9.77 years. A majority of the sample indicated speaking English in the home (79.5%), and all child participants were fluent English speakers. | First Nations, Inuit, and Metis | Anxiety reduction |
| Mota 2012 | Peer-reviewed | Examine associations of individual, friend or family, and community or tribe factors with suicidality in a representative on-reserve sample of more than 1100 First Nations adolescents. | Observational | Manitoba | Suicide | Adolescents aged 12 to 17 years | First Nations | Suicidality, depression, substance use, languages, cultural activities, religious beliefs, socioeconomic disparity, residential schooling, previous suicide by friend or family, traumatic life events, community of residence |
| Njeze 2020 | Peer-reviewed | Contributes to a growing body of literature examining aspects of resilience and wellness among urban Indigenous youth, and critically engage with and expand intersectionality theory to consider—in addition to overlapping forms of oppression or marginalization—how strengths based and positive individual, social, and cultural factors can also intersect and coalesce. | Case study | Saskatchewan, Canada | General mental health and well-being | 28 youth from nêhiyaw (Plains Cree; n = 19), Dene (n = 2), and Métis (n = 7) backgrounds between the ages of 16 and 24 years (12 male and 16 female). | Cree, Dene, Metis | Perceptions, experiences related to resiliency |
| Paul 2022 | Peer-reviewed | Examine associations of bullying and cyberbullying with psychological distress in a nationally representative sample of youth aged 12 to 17 living in their First Nations communities in Canada. | Observational | Canada (National) | Anxiety and depression | Living in First Nations communities across Canada. n=4,968 Male:n=2,514 Female:n=2,454 | First Nations | Community belonging; participation in community cultural events; importance of traditional cultural events; psychological distress |
| PetrasekMacDonald 2015 | Peer-reviewed | Examine protective factors from a youth perspective and could offer new pathways to resilience and adaptive capacity within the context of climate change. | Case study | Nunatsiavut, Labrador | General mental health and well-being | 17 youths, aged 15-25, between two and five males and females, participated in an interview | Inuit | Youth's perspectives on protective factors that enhance mental health and well-being, focusing on factors that are challenged by climate change. |
| Rawana 2012 | Peer-reviewed | Identify protective factors related to the alcohol use trajectories from early adolescence to emerging adulthood among off-reserve Canadian Aboriginal youth. | Observational | Yukon or Northwest Territories, individuals living in institutions, and individuals living on reserves | Substance use | 330 participants, 50.3% male aged 12–23 | Inuit Metis | Alcohol use, participation in activities, volunteer participation, attendance of religious services, positive peer relationships, prosocial behaviours, self-esteem and optimism. |
| Rawana 2015 | Peer-reviewed | Describe the development and preliminary evaluation of a peer mentorship program for Aboriginal students attending a large urban Canadian university | Mixed methods | N/A | General mental health and well-being | Of the eight participants, one was a first year student, two were second year students, four were fourth year students, and one was a graduate student. | First Nations, Metis and Inuit | Program evaluation - perceptions and experience |
| Ritchie 2014 | Peer-reviewed | To evaluate the impact of a culturally relevant Outdoor Adventure Leadership Experience (OALE) on the resilience and well-being of adolescents from Wikwemikong Unceded Indian Reserve in north-eastern Ontario.” | Mixed methods | Ontario, Canada | General mental health and well-being | 73 participants ranged in age from 11.9 to 18.7 years with a mean age of 14.6, and 64.4% (n=38) were male. Less than half (42.4%; n=25) of all participants lived in a family situation with both parents. No participants self-identified as non-Aboriginal. | First Nations, Inuit, and Metis. | Resilience, other exploratory measures including health and well-being: mental, physical, emotion, self-esteem, satisfaction with life  spiritual values, social support, community values |
| Ritchie 2015 | Peer-reviewed | Qualitative evaluation of a larger collaborative project to develop, implement and evaluate an outdoor adventure leadership experience (OALE) for youth ages 12–18 from Wikwemikong Unceded Indian Reserve in northeastern Ontario, Canada | Qualitative | Wikwemikong Unceded Indian Reserve, northeastern Ontario, Canada | General mental health and well-being | A total of 43 youth, ages 11.9–18.7 years. The mean age of the youth was 14.7 years, and there were 16 (37%) female youth participants | First Nations | Connecting with good life, connecting with creation, connecting with self, viewing through indigenous lens |
| Schick 2022 | Peer-reviewed | Examine subjective happiness as a moderator of the link between alcohol expectancies and alcohol use (i.e., to examine whether the effect of alcohol expectancies on alcohol use is conditional on one’s level of subjective happiness) | Observational | Ontario | Substance use | Participants ranged in age from 11 to 18 (M=14.58, SD=2.15), were 50.0% female (n=53) and 50% male (n=53). | First Nations | Alcohol expectancies, subjective happiness, alcohol use, analytic strategy |
| Scott 1988 | Peer-reviewed | Investigate the effect of physical training on native adolescents’ self-evaluation and substance use patterns. | Observational | Maniwaki, Quebec | Substance | 30 students (16 females, 14 males) Age range 12-18, grades 7-11 from the River Desert Community | First Nations | Drug-related attitudes and behaviour, fitness, self esteem |
| Snowshoe 2017 | Peer-reviewed | Examine the relationships between culture and mental health among FN youth using a brief version of the newly developed Cultural Connectedness Scale | Observational | Saskatchewan and Southwestern Ontario | General mental health and well-being | 290 FN, Me´tis, and Inuit youth (140 male, 140 female; 10 unspecified) enrolled in grades seven through 12 from Saskatchewan (n = 153) and Southwestern Ontario (n = 137) in Canada. Approximately 68 percent of respondents reported living on-reserve. Respondents ranged in age from 11 to 24 years of age (M = 14.4; SD = 2.4), with approximately 90 percent being age 18 or under. | First Nations, Inuit, and Metis. | Cultural connectedness, stressful life events, self-efficacy, sense of self in the present and in the future, school connectedness, life satisfaction |
| Spillane 2020 | Peer-reviewed | Investigate risk and protective factors associated with substance use in one group of First Nation adolescents. | Participatory Action Research | Eastern Canada | Substance use | A total of 15 (5: male and 10: female) FN adolescents participated in the focus groups, and 11 of these adolescents participated in the individual interviews. | First Nations | Perceptions about substance use |
| Spillane, Schick, Nalven et al., 2021 | Peer-reviewed | Examine the association of importance and availability of the three categories of Competing Life Reinforcers CLRs (i.e., cultural, social, and extracurricular activities) with alcohol and marijuana use behaviours to test the applicability of Behavioral Theories of Choice BTC to a group of adolescents living in rural Indigenous reserve communities in Canada. | Observational | Rural areas of Eastern Canada | Substance use | 106 First Nation adolescents from Indigenous communities located in rural areas of eastern Canada. Participants ranged in age from 11 to 18 years and were in grades 6 through 12 with four participants no longer attending school.  Half the sample (50.0%) identified as female, and all reported that they were a member of a First Nation group and lived within reserve communities. | First Nations | Marijuana use, alcohol use, reinforcers |
| Spillane, Schick, Goldstein et al., 2021 | Peer-reviewed | Examine how self-compassion relates to alcohol use, alcohol-related problems, and AUD risk in a sample of First Nation adolescents. | Observational | eastern Canada | Substance use | 106 First Nation adolescents from Indigenous communities located in Eastern Canada, ranged in age from 11 to 18 years (M=14.6, SD=2.2), were 50.0% female, and all self-reported that they were a member of a First Nation group and living on reserve. | First Nations | Alcohol use, alcohol-related problems, and self-compassion. |
| Thomas 2022 | Peer-reviewed | Investigate how COVID-19 has affected the wellness of a group of Inuit youth leaders in Nunavut in the context of their involvement with an ongoing mental health research initiative, the Making I-SPARX Fly in Nunavut [I-SPARX] project. The study had three goals: (1) to understand how the pandemic has affected I-SPARX leaders’ perceived involvement in the I-SPARX Project; (2) to build knowledge around how the pandemic has impacted the daily life and wellbeing of youth in Nunavummiut communities; and (3) to acquire a culturally specific understanding of their coping mechanisms and resilience strategies through the lens of Inuit Qaujimajatuqangit (IQ). | Qualitative | Nunavut  They came from four communities in Nunavut: Cambridge Bay, Baker Lake, Pond Inlet, and Cape Dorset | General mental health and well-being | 9 Inuit youth who participated in this study were between the ages of 16 and 22 (M = 18, SD = 1.94). Three participants identified as female and six as male. | Inuit | Experience in I-SPARX, their life during the pandemic, and their coping strategies. |
| Turner 2001 | Grey lit | Explore factors (eg, depression, substance use, parental support. self appraisal) that predict or prevent a particular maladaptive behavior--propensity for suicide attempt--among a large sample of Canadian minority adolescents, specifically British Columbia First Nations youth; determine the extent to which the selected factors contribute to high or low propensity for suicide, and whether these findings differ from those for non-First Nations youth. | Observational | British Columbia | Suicide, substance use, depression, emotional stress | The mean age of the First Nations adolescents was 15.3 1 (SD = 1.87), and a roughly equal representation of males and females in the study. | First Nations | Propensity for suicide and contributing factors |
| Walls 2007 | Grey lit | Examine correlates of Indigenous adolescent suicidality from within a community based participatory research (CBPR) framework. | Mixed methods | Calgary | Suicide | A majority (97%) of the 746 youths interviewed were 10 - 12 years old | First Nations | Suicidal ideation and behaviour, alcohol use, depressive symptoms, youth delinquency, youth anger, self esteem |
| Wood 2020 | Peer-Reviewed | Build upon the following question: What relational and physical spaces and places support youth civic and cultural engagement? Explore both the existing and required resources in communities that supported youth civic and cultural connection. | Participatory Action Research | “The research site is a fly-in community on Labrador’s North Coast” | General mental health and well-being | 8 youth of which five were young women (aged 15–16 at the start of the study) and three were young men (aged 13, 16 and 17). | Inuit | Existing and required resources in communities that supported youth civic and cultural connection |
| Zahradnik 2010 | Peer-reviewed | Rest the hypotheses that exposure to violence among a sample of Mi’kmaq students would positively correlate with PTSD symptoms, but that resilience would negatively correlate with PTSD; Ivestigate whether resilience moderates the relationships between exposure to violence and PTSD | Observational | Nova Scotia | Post-traumatic stress disorder, trauma | 126 students of which 72 were female, 53 were male, one gender not indicated). Of the 126 students, the vast majority of the students (78.0%) were 16 years of age or older, the age at which according to Nova Scotian Law , a child can choose whether or not he or she wishes to report a case of abuse where he or she alone was the victim. Approximately one third of the students (29.4%) indicated that they were 18 years of age or older. The self reported education level achieved ranged from grades 8–12 (Mean=10). | Mi'kmaq | Resilience, violence, post-traumatic stress disorder |
