## Supplementary File 4 for "Resilience and Protective Factors for Mental Health among Indigenous Youth in Canada: A Scoping Review"

| Supplementary File 4. Definitions | | |
| --- | --- | --- |
| Author Year | Definition of resilience and/or protective factors identified in included studies | Level(s) |
| Ames 2015 | The developmental psychopathology perspective presumes that various factors work dynamically to negate negative mental health consequences. | Individual |
| Back Feathers of Hope 2014 | Commonly used to talk about the ability a person has to “bounce back,” adapt or cope with stress when they experience problems or major challenges in their lives | Individual |
| Dell 2011 | A balance between individual strategies of coping with adversity and the availability of community support. | Individual + Community |
| Fanian 2015 | Both the capacity of individuals to navigate their way to the psychological, social, cultural, and physical resources that sustain their well-being, and their capacity individually and collectively to negotiate for these resources to be provided in culturally meaningful ways. | Individual + Community |
| Filbert 2010 | Resilience, defined as average or above-average functioning in the face of serious threats to development. | Individual |
| Filbert 2014 | Resilience: Positive adaptation in the face of serious threats to development Protective factors: Assets or resources refer to direct predictors of positive outcomes, and those assets that appear to function under high-risk or high-adversity conditions. | Individual + Community |
| Hatala 2017 | A pattern of positive adaptation in the midst of or following significant stress, adversity, or risk | Individual |
| Hirsch 2016 | Individual’s ability to overcome stress and adversity | Individual |
| Isaak 2020 | Displaying substantial strength despite perceived or experienced barrier | Individual |
| Kral 2014 | Less as an individual trait but rather through the social context as a process. | Community |
| Liebenberg 2022 | Resilience is understood as the ability to do well, or better than expected, despite adversity | Individual |
| Njeze 2020 | Reduced vulnerability to environmental risk experiences, the overcoming of stress or adversity, or a relatively good outcome despite acute distress. Historically grounded in their persistence, resourcefulness, and adaptability to the unpredictable Arctic environment, as well as their ability to navigate uncertain environmental challenges. | Individual |
| PetrasekMacDonald 2015 | The balance between maintaining cultural traditions, beliefs, and identity and being flexible in responding to changing social, cultural, or environmental conditions. | Individual + Community |
| Rawana 2012 | Protective factors work to negate or confer against alcohol abuse and dependence disorder diagnoses or alcohol use related problem | N/A |
| Rawana 2015 | Resilience is defined as protective factors that allow for positive adjustment and coping toward stressors | N/A |
| Ritchie 2014 | The ability to successfully cope with change or misfortune | Individual |
| Ritchie 2015 | The ability to successfully cope with change and misfortune | Individual |
| Thomas 2022 | Resilience can be defined as the ability to positively cope, find hope, and foster constructive outcomes in contexts of adversity. | Individual |
| Turner 2001 | Resilience has generally been defined as the adaptive competence and absence of psychiatry disorders despite the presence of significant psychosocial stress, trauma, illness, or loss; "as good developmental outcomes despite high-risk status, sustained competence under stress, and recovery from trauma"; when positive outcomes are evidenced in the face of being at significant risk for developing problems; and when "one regains functioning following adversity to the level of adaptation and competence that characterized the individual prior to the pre-stress period. | Individual |
| Wood 2020 | An interactive process, integrating individual resources together with social and physical resources. | Individual + Community |
| Zahradnik 2010 | Ability to thrive in the presence of adversity | Individual |
