## Supplementary File 5 for "Resilience and Protective Factors for Mental Health among Indigenous Youth in Canada: A Scoping Review"

| **Author, Year** | **Level of involvement** | **Involved members** |
| --- | --- | --- |
| Adams 2015 | Consult | Elder, Indigenous community members/leader/workers, decision makers/ government |
| Ames 2015 | NR | NR |
| Ansloos, J., Dent, E. 2021 | Consult | NR |
| Arato-Bollivar 2005 | NR | NR |
| Baydala 2014 | Empower | Elder, Indigenous community members/leader/workers |
| Bohr 2016 | NR | Decision makers/ government |
| Chandler 1998 | Involve | NR |
| Crooks 2010 | Collaborate | Community members/workers not specified ,youth, school/program administrators |
| Crooks & Dunlop 2017 | Collaborate | Community members/workers not specified ,youth |
| Crooks 2017 | NR | NR |
| Dell 2011 | NR | NR |
| Dell and Hopkins 2011 | Consult | NR |
| Dunlop 2016 | NR | NR |
| Eggertson 2013 | Inform | Decision makers government, Indigenous community members/leader/workers |
| Fanian 2015 | Empower | NR |
| Feathers of Hope 2014 | empower | Indigenous community, decision makers |
| First Nations Information Governance Centre 2014 | Collaborate | Elder, family, community not specified, school/program |
| First Nations Information Governance Centre 2021 | Collaborate | Community members/workers not specified |
| Filbert 2010 | NR | NR |
| Filbert 2014 | Collaborate | Youth, family, Healthcare provider |
| Flanagan 2011 | Involve | Elder, Indigenous community members/leader/workers, school/program administrators |
| Fraser 2015 | NR | NR |
| Gfellner 2016 | Collaborate | Indigenous community members/leader/workers, community members/workers not specified |
| Goldstein 2021 | Consult | Indigenous community members/leader/workers, family |
| Hackett 2016 | Involve | Community members/workers not specified |
| Hallett 2007 | NR | NR |
| Hammond 2000 | NR | Healthcare provider |
| Hatala 2017 | Involve | Elder, community members/workers not specified, youth, family |
| Hirsch 2016 | Collaborate | Community members/workers not specified ,youth, school/program administrators |
| Hutt-MacLeod 2019 | Involve | Healthcare provider, youth, family, community members/leader/workers not specified |
| Isaak 2020 | Collaborate | Indigenous community members/leader/workers |
| Ivanich 2020 | Empower | Elder, community members/workers not specified ,youth, family, school/program administrators, healthcare provider |
| Janelle 2009 | Collaborate | Indigenous community members/leader/workers |
| Katapally 2020 | Collaborate | Youth, school/program administrators, community members not specified |
| Kirmayer 1998 | NR | NR |
| Kral 2014 | Collaborate | Elder, community members/workers not specified ,youth |
| Liebenberg 2022 | Collaborate | Indigenous community members/leader/workers, youth, healthcare provider |
| Linds 2019 | Empower | Indigenous community members/leader/workers, community members/workers not specified, youth |
| Litwin 2023 | Involve | Healthcare provider, decision makers/government |
| Lys 2018 | Involve | Youth, school/program administrators |
| MairTiessen 2008 | Consult | Indigenous community members/leader/workers, community members/workers not specified, youth, healthcare provider |
| Miller 2011 | Inform | NR |
| Mota 2012 | NR | NR |
| Njeze 2020 | Collaborate | Indigenous elder, youth, family |
| Paul 2022 | NR | NR |
| Petrasek MacDonald 2015 | Consult | Elder, community members/workers not specified |
| Rawana 2012 | NR | Youth |
| Rawana 2015 | Involve | Elder, Indigenous community members/leader/workers, community members/workers not specified |
| Ritchie 2014 | Consult | Community members/workers not specified, youth |
| Ritchie 2015 | Collaborate | Community members/workers not specified |
| Schick 2022 | NR | NR |
| Scott 1988 | NR | NR |
| Snowshoe 2017 | Involve | Community members/workers not specified |
| Spillane 2020 | Collaborate | Community members/workers not specified, youth, family |
| Spillane, Schick, Nalven et al., 2021 | NR | NR |
| Spillane, Schick, Goldstein et al., 2021 | NR | NR |
| Thomas 2022 | Collaborate | Youth |
| Turner 2001 | NR | NR |
| Walls 2007 | NR | Elder, community members not specified, program administrator/school |
| Wood 2020 | Collaborate | Youth |
| Zahradnik 2010 | NR | NR |
