## Supplementary File 6 for "Resilience and Protective Factors for Mental Health among Indigenous Youth in Canada: A Scoping Review"

**Hwayeon Danielle Shin (she/her)** is of Korean ancestry, who immigrated to the traditional and unceded territory of the Mi’kmaq people during her youth. She is a nurse and is presently situated in the traditional territory encompassing several nations, including the Mississaugas of the Credit, the Anishnabeg, the Chippewa, the Haudenosaunee, and the Wendat peoples, which is recognized as Toronto.

**Leah Carrier (she/her)** is a Two-Spirit mixed Indigenous (Niitsitapi) and European-Canadian settler nurse currently based in Mi’kma’ki, the traditional and unceded territory of the Mi’kmaq people. She was raised by her settler Canadian mother in St’at’imc territory and is on a reconnecting journey with her Indigenous heritage.
